# Bibliometric and visualized mapping: two decades of lipidomics, with special focus on pregancy and women

**DOI:** 10.1101/2023.09.07.23295179

**Authors:** Lin Zhang, Ying Zhou, Jiashun Zhou

## Abstract

To perform a bibliometric visualization in lipidomics-related research with two decades. The primary data was retrieved from the Web of Science, three sotwares (VOSviewer, CiteSpace, and R) provided an overview of this field. The countries, institutions, authors, key terms, and keywords were tracked and corresponding mapping was generated. From January 1st in 2001 to March 21th in 2022, 45,325 authors from 234 organizations in 101 countries published 7,338 publications in 382 journals were found. *Journal of Lipid Research* was the most productive (284 publications) and highly cited journal (18,293 citations). We clustered four keywords themes. The niche theme were shotgun lipidomics, tandem mass-spectrometry, and electrospray-ionization. The motor theme were expression, diseases, and inflammation. The emerging or decling theme were identification, mass-spectrometry, and fatty acids.The basic theme were metabolism, cell, and plasma. Though eight categories the lipid were classified, the keywords showed two of which were got more attention for research, fatty acyls and glycerophospholipids. The top 3 lipidomics-favoured diseases were insulin resistance, obesity, and Alzheimer’s disease. The top 3 lipidomics-favoured tissue was plasma, brain, and adipose tissue. Burst citations show “women” and “pregnancy” with the strength of 8.91 and 7.1, both topics may be a potential hotspot in the future.

## 1. Introduction

Lipidomics aims to systematically analyze the alteration in lipid composition and expression of various organisms, which can effectively reveal the changes and functions of various biological processes, such as energy metabolism (Moellering and Cravatt, 2013), inflammation, and immunity (Que et al., 2018). Though put forward earlier, the concept of lipidomics was widely accepted because of Prof. Xian-Lin Han (Han and Gross, 2003) until 2003, it has received increasing attention in numerous fields in the past two decades. As a crucial branch of metabolomics, lipidomics was certified to play an indispensable role to maintain the balance of the individual. Especially in the health field, lipidomics has been widely applied in the study of diverse diseases, such as kidney disease (Zhang et al., 2016), immunological diseases (Zeng et al., 2017), metabolic disease (Overgaard et al., 2018; Gurgul-Convey, 2020), and tumors (Umezu-Goto et al., 2004; Bougnoux et al., 2006; Sabbadini, 2006). Furthermore, lipidomics was involved in other files, such as gut microbiota (Velagapudi et al., 2010; Farrokhi et al., 2013), food safety (Liu et al., 2022; Sun et al., 2022), and pathogenic microorganisms (Hines and Xu, 2019; Danne-Rasche et al., 2020; Péter et al., 2021).

According to the LIPID MAPS (http://www.lipidmaps.org), the lipids (current 47,449 types) can mainly classify into the following eight categories, including fatty acyls, glycerolipids, glycerophospholipids, sphingolipids, sterol lipids, saccharolipids, prenol lipids, and polyketides. Furthermore, with or without unsaturated hydrocarbon chains, lipids can be divided into unsaturated or saturated lipids (Williams et al., 1966). Because of the multiple-type of structures and increasing publications of lipidomics, it is hardly to generate a panoramic assessment of this field. A manual compilation and systematic review of all the publications in this field would be, if not impossible, time-consuming. With the surged research and complicated composition of lipids during the past two decades, it is necessary to portray the character of these articles to further summarize the trends at present and possible concerns in the future.

Bibliometric analysis has been used to process the above-mentioned problems in other disciplines. Most bibliometric computational and visual analytic approaches are automated and thus suitable for analyzing tremendous amounts of literature on broad and various topics. Bibliometrics has been adopted to assess the research status in diverse fields, such as metabolomics (Robertson et al., 2011) and mass spectrometry (Waaijer and Palmblad, 2015). Unlike traditional citation counts, bibliometrics considers connections among literatures, enabling the identification of intellectual structures and emerging trends (Raghupathi and Nerur, 2008). Bibliometrics identifies relevant nodes and extracts useful information from a large amount of information (Zhang et al., 2018; Qi et al., 2019; Tan et al., 2021) and promotes researchers a better understanding of the emerging trends and knowledge structure in the temporal cross-section of a research field (Dong et al., 2020; Liang et al., 2020; Lin et al., 2020). Although a lot of bibliometric software is used, including CiteSpace (Liang et al., 2017; Chen et al., 2019), VOSviewer (van Eck and Waltman, 2010), bibExcel (Shamsi et al., 2020), Science of Science (Wu et al., 2022), and HistCite (Li et al., 2022), CiteSpace and VOSviewer are the most frequently applied software. CiteSpace (Chen, 2020) is a freely available Java application designed by Professor Chao-Mei Chen that aims to detect new trends and mutations in scientific literatures. VOSviewer (van Eck and Waltman, 2010) was created by the Centre for Science and Technology Studies of Leiden University and provided visually better pictures compared with CiteSpace. Bibliometrics usually focuses not only on the current trends, but also on the hotspots in the future (Lim et al., 2021). Therefore. bibliometrics is quite suitable for objective academic evaluation that is conducive to evaluating the academic impact of lipidomics.

This study aims to integrate bibliometric approaches to analyze the literatures on lipidomics with the deadline of March 21th in 2022. To the best of our knowledge, this is the first bibliometric analysis in this field. Our findings will offer a comprehensive overview of this field and highlight its intellectual structures with emerging trends. The revelation of journal preferences may help researchers choose appropriate journals. Furthermore, highlighting the strengths of the institutions in this field may help researchers make career-related decisions. Moreover, the discovery of emerging trends may help the researchers determine the direction of their studies.

## 2. Materials and methods

### 2.1. Bibliographic records

Briefly, the publication data were performed on March 21th in 2022, and downloaded as a “Plain text file” with full record and cited reference from the Web of Science Core Collection (WOScc). The retrieval strategies and data collection process need to meet the following criteria:

1. the search strategy was guided by the TS (“topic” including title, abstract, author’s keywords, and keywords Plus) as (((TS=(Lipidomics)) OR TS=(Lipidomic)) OR TS=(Lipidome)) OR TS=(Lipidomes);
2. the document type was “article”;
3. the publication deadline was March 21, 2022;
4. the following information was collected: publication, authors, Countries, institutions, journals, keywords, and citations.

### 2.2. Bibliographic analysis

With the basement of Java (version 11.0.12), CiteSpace (version 6.0 R1) was utilized for the data analysis. In CiteSpace, the “time-slicing” was confined to 2001-2022 with the “years per slice” into “1”. Furthermore, the node types were classified into Country, institution, source, reference, author, and keywords. The “top N per slice” was set as “50” which means the 50 documents with the highest cited frequency were selected for each “time slicing”. In addition, we chose “pathfinder” for the network analysis. To better indicate the correlation of the above project, VOSviewer (version1.6.17) was performed for visualization. Finally, R (version 4.1.2) was used to summarize the frequency with the package *tableone* and portrayed the world distribution of research with packages of *ggplot2*, *ggmap*, and *maps*. And R package *biblimotrix* was applied into clusterd, thematic evalution, and trend topics analysis.

### 2.3. Research ethics

Bibliographic information was searched and downloaded from WOScc (publicly available database). The extraction data of this research did not involve interaction with human subjects or animals. Thus, our study is immune to ethical issues. No approval from an ethics committee was required.

## 3. Result

### 3.1. Annual global publication outputs of lipidomics

According to the searching strategy, we retrieved 9,311 primary publications data through the database. With the filter criteria of inclusion and exclusion, 7,338 articles were chosen for the following analysis at last (**Fig. 1**).

**Fig. 1.**
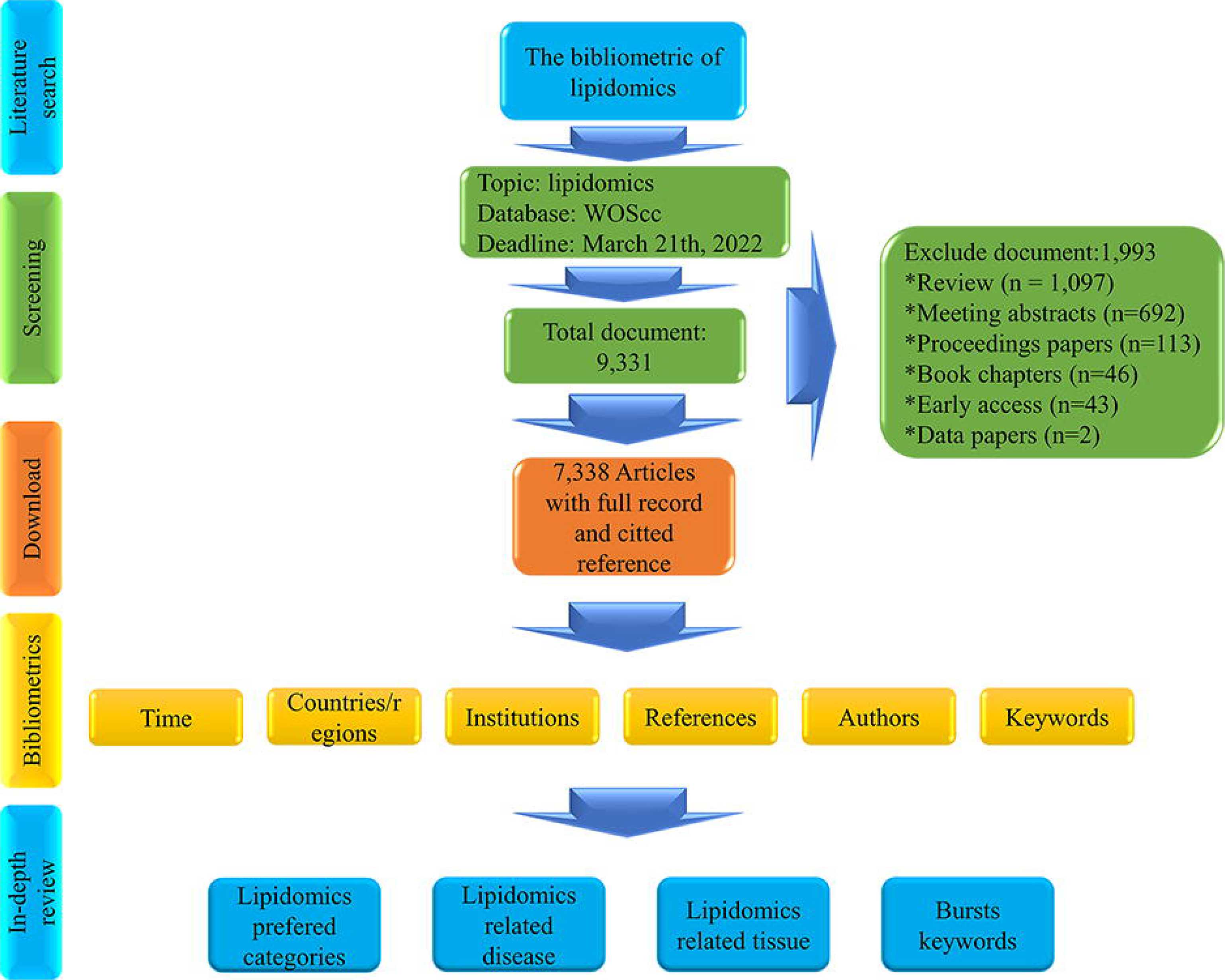
The workflow of the analysis steps.

The annual distribution (**Fig. 2**) indicated that lipidomics-related publications were surging during the past two decades. Because of the deadline time constraints, only 176 articles were calculated in 2022, the inclusion of articles in 2022 is not complete and the time distribution did not consider 2022. Although the concept “lipidomics” was widely recognized until 2003, still put forward a little earlier. The total time distribution can be divided into two segments. One is the downturn period from 2001 to 2004 with at most 11 articles. Another is the increasing period from 2005 to 2021with 44 articles to 1,437 articles, respectively. A trend line (**Fig. 2**), used to evaluate the imitative effect, indicated that when excluding the incomplete 2022 publication, the trend was exponential from 2001 to 2021 and R^2^ is higher than 0.97.

**Fig. 2.**
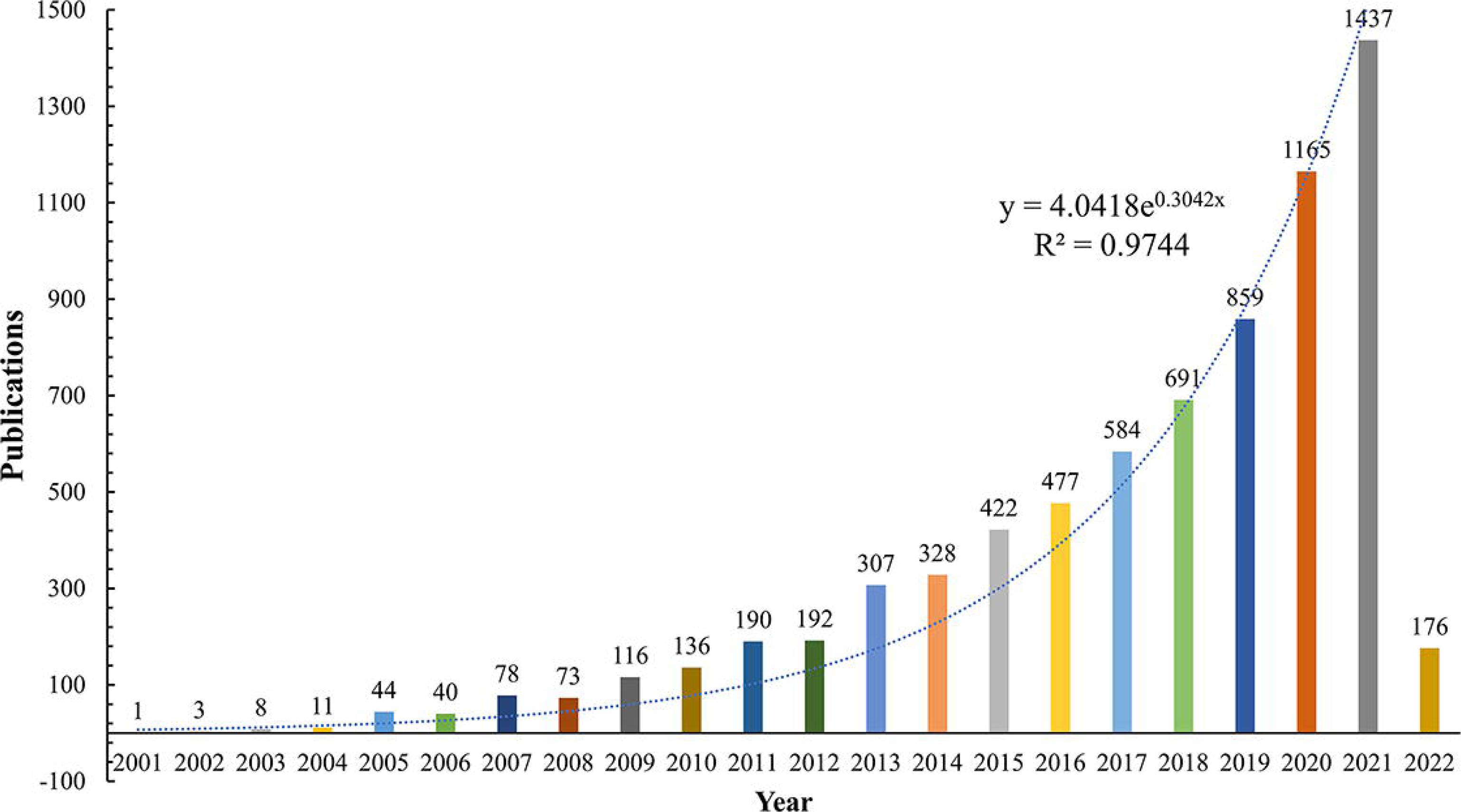
The time distribution between January 1, 2001, to March 21, 2022.

### 3.2. Distribution of Countries/regions and institutions

A total of 101 Countries were involved in the lipidomics with 1 to 2,577 publication articles (**Table S1**), and 38 Countries published more than 20 articles (**Fig. 3A**). The USA (2,577 publications with 110,883 citations) was the most productive Country in the world, followed by China (1,517 publications with 20,815 citations), Germany (893 publications with 31,478 citations), England (574 publications with 23,754 citations), France(441 publications with 12,763 citations), Italy(409 publications with 12,282 citations), and Australia(399 publications with 12,697 citations).

**Fig. 3.**
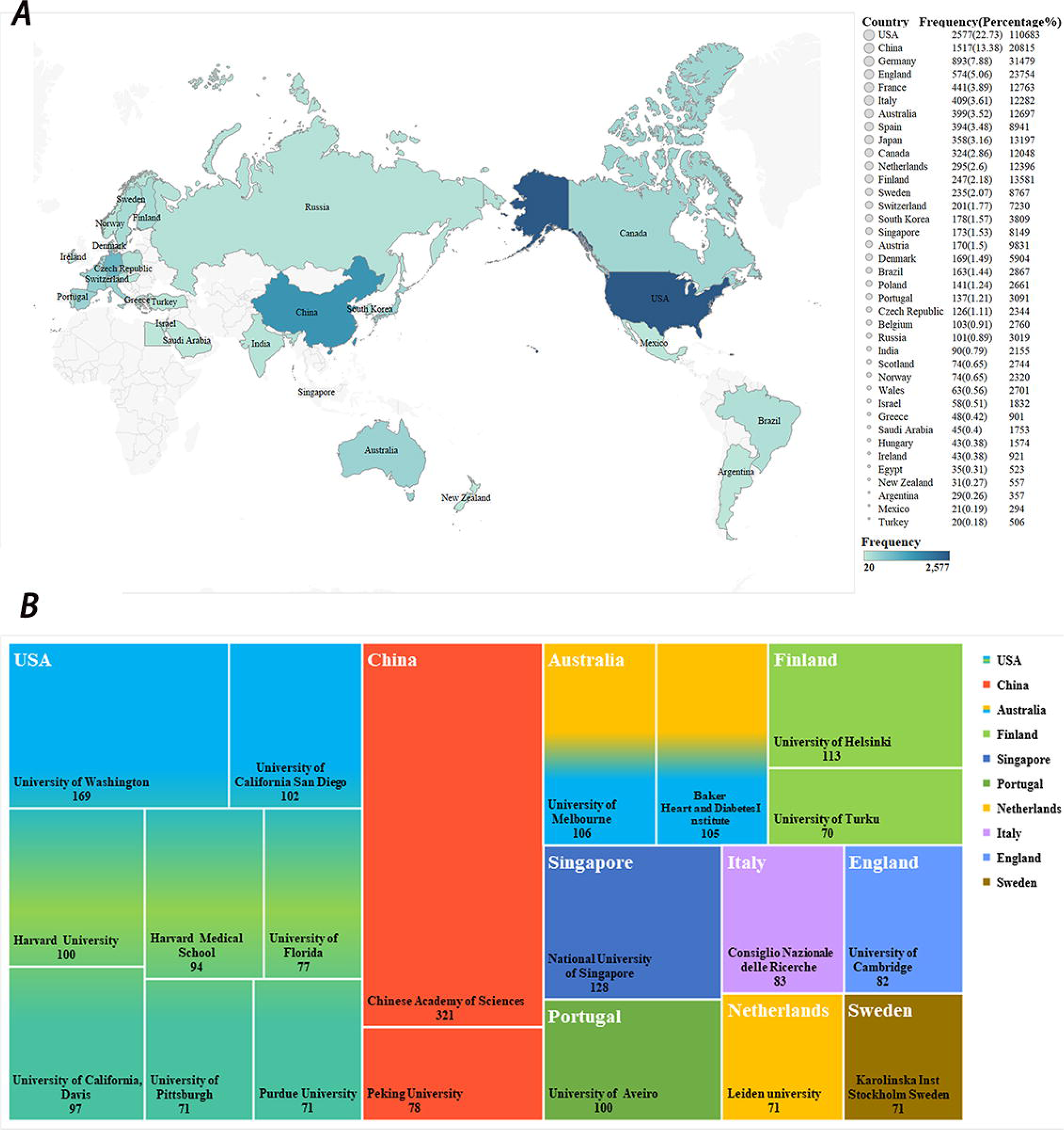
The distribution of Countries/regions and institutions. A) The top 38 Countries/regions with more than 20 publications and 14 Countries/regions cannot show on the map because of the cartographic limitation. B) The top 20 productive institutions around the world.

To display the national distribution with the various period, we summarize the top 10 Countries in four periods (**Table 1**), including 2001-2007, 2008-2012, 2013-2017, and 2018-2022, respectively. In the first period (2001-2007), the USA (*n* = 113) was the most productive Country in lipidomics with a great discrepancy compared with followed England (*n* = 24) and Germany (*n* = 18). In the second period (2008-2012), the USA (*n* = 311) was still top 1, following Germany (*n* = 92) and Finland (*n* = 55). In the third period (2013-2017), although the USA (*n* = 300) still was top 1, China (*n* = 300) replaced Germany (*n* = 273) in second place. In the fourth period (2018-2022), the USA (*n* = 1,368) and China (*n* = 1,162) were still the top 1 and 2, however, the difference was quite small. The top 3 Countries (USA, China, and Germany) contribute 43.99% of total. Although China had a late beginning on lipidomics research, it displayed a surging trend during the four periods. As for the citations, Chinese publications only ranked at fourth, which might be caused by the late beginning.

**Table 1.**
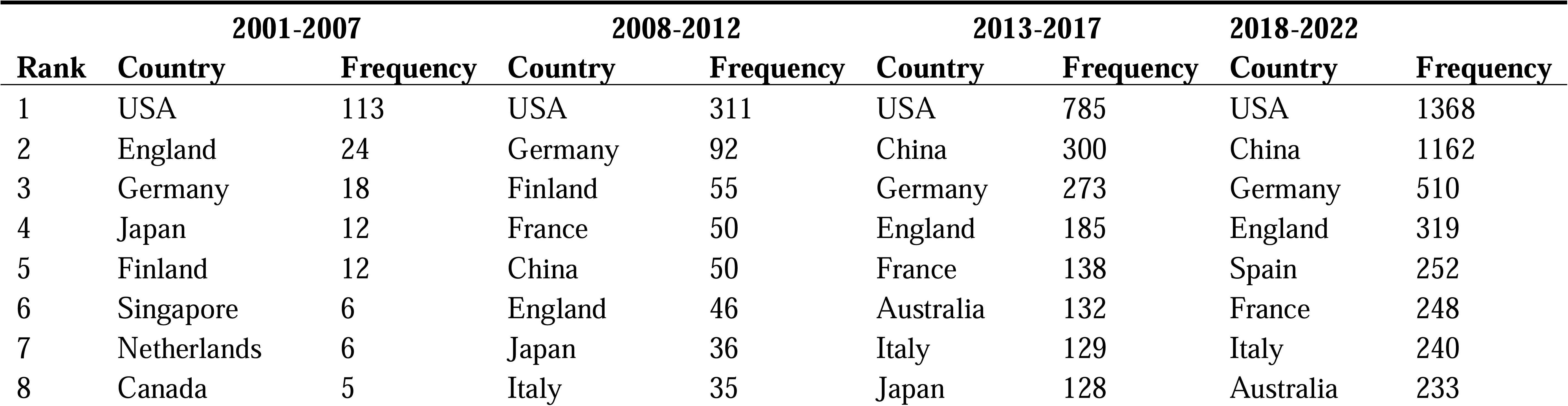
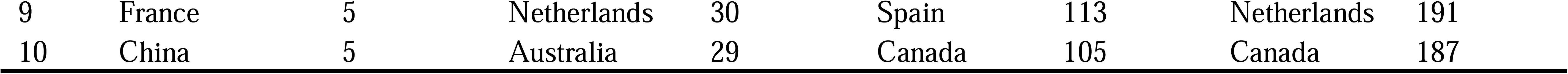
The top 10 Countries/regions of distribution in four periods.

A total of 234 institutions, which published more than 10 lipidomics-related articles (**Table S2**), were dominantly derived from the USA, China, and Germany. The articles of these institutions were mainly contributed by universities, followed by hospitals and research laboratories.

In the top 20 institutions with the derived Countries (**Fig. 3B**), USA and China were the majority Countries, followed by Australia, Singapore, Finland, and Portugal. The citations of Harvard University (*n* = 10,952) were top 1, however the publications (*n* = 100) were quite less, which emphasized the academic status and influence of Harvard University. Conversely, the publications of the Chinese Academy of Sciences (*n* = 321) ranked at 1^st^, while the citations (*n* = 5,699) were only at 7^th^, which might be caused by the late beginning of this aera in China.

To optimally dynamic demonstrate the institutions with diverse periods, we summarize the top 5 institutions in four periods (**Table 2**) respectively, including 2001-2007, 2008-2012, 2013-2017, and 2018-2022. In the first period (2001-2007), the top 3 institutions were all from the USA, including University of Washington, Harvard University, and Brigham and Women’s Hospital. In the second period (2008-2012), the first institution was still the University of Washington, while both the second and third were derived from Finland (VTT Technical Research Centre of Finland and The University of Helsinki). In the third period (2013-2017), the Chinese Academy of Sciences had turned to be the top 1, the University of Washington fell to third position, and the Heart and Diabetes Institute of Australia was the second. In the fourth period (2018-2022), the Chinese Academy of Sciences also ranked at the top 1, followed by Harvard Medical School and the University of Melbourne.

**Table 2.** The top 5 institutions in four periods.

| 2001-2007 |  |  |  | 2008-2012 |  |  |
| --- | --- | --- | --- | --- | --- | --- |
| Rank | Institution | Country | Frequency | Institution | Country | Frequency |
| 1 | University of Washington | USA | 30 | University of Washington | USA | 41 |
| 2 | Harvard University | USA | 11 | VTT Technical Research Centre of Finland | Finland | 38 |
| 3 | Brigham and Women's Hospital | USA | 7 | University of Helsinki | Finland | 29 |
| 4 | University of Helsinki | Finland | 7 | Harvard University | USA | 27 |
| 5 | Kansas State University | USA | 6 | National University of Singapore | Singapore | 25 |
| 2013-2017 |  |  |  | 2018-2022 |  |  |
| Rank | Institution | Country | Frequency | Institution | Country | Frequency |
| 1 | Chinese Academy of Sciences | China | 98 | Chinese Academy of Sciences | China | 211 |
| 2 | Baker Heart and Diabetes Institute | Australia | 53 | Harvard Medical School | USA | 76 |
| 3 | University of Washington | USA | 47 | University of Melbourne | Australia | 71 |
| 4 | National University of Singapore | Singapore | 44 | University of Florida | USA | 66 |
| 5 | Harvard University | USA | 38 | University of Aveiro | Portugal | 65 |

### 3.3. Distribution of Source Journals and top 15 high co-cited references

The retrieved articles on lipidomics were published in 382 journals. We summarize the top 10 journals (**Table 3**) which published the most lipidomics-related articles and account for 26.5% (1,942/7,338) of all the publications. *Journal of Lipid Research* was the most productive journal (284 publications) and the most highly cited journal (18,293 citations), followed by *Scientific Reports* with 262 publications and *Analytical Chemistry* with 241 publications.

**Table 3.**
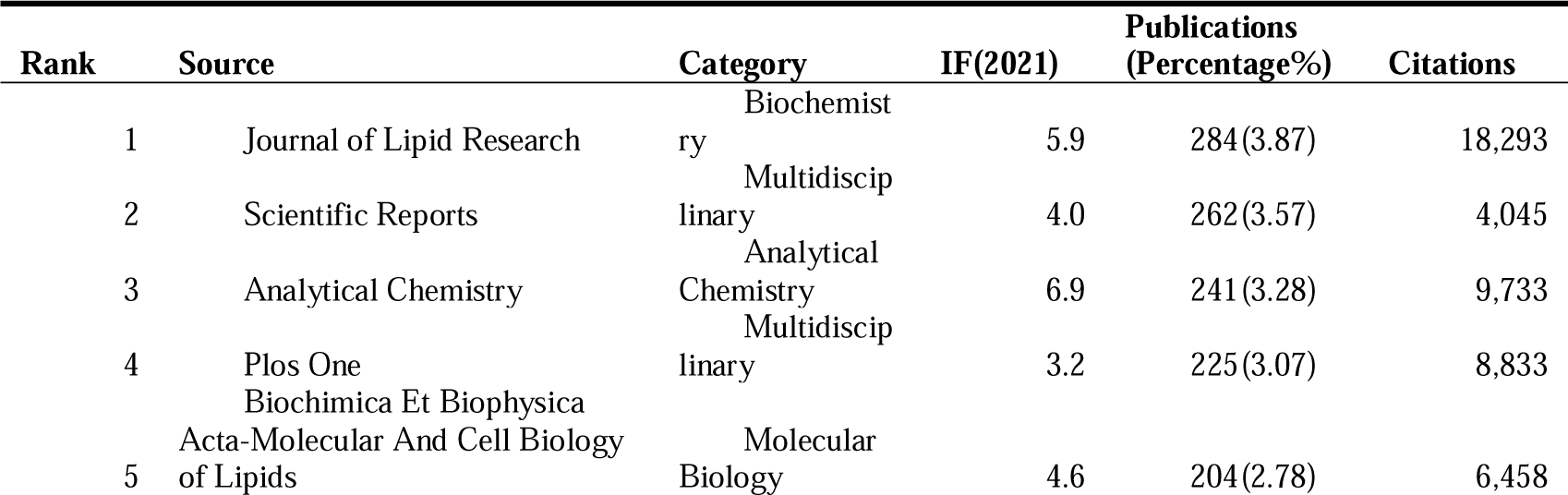
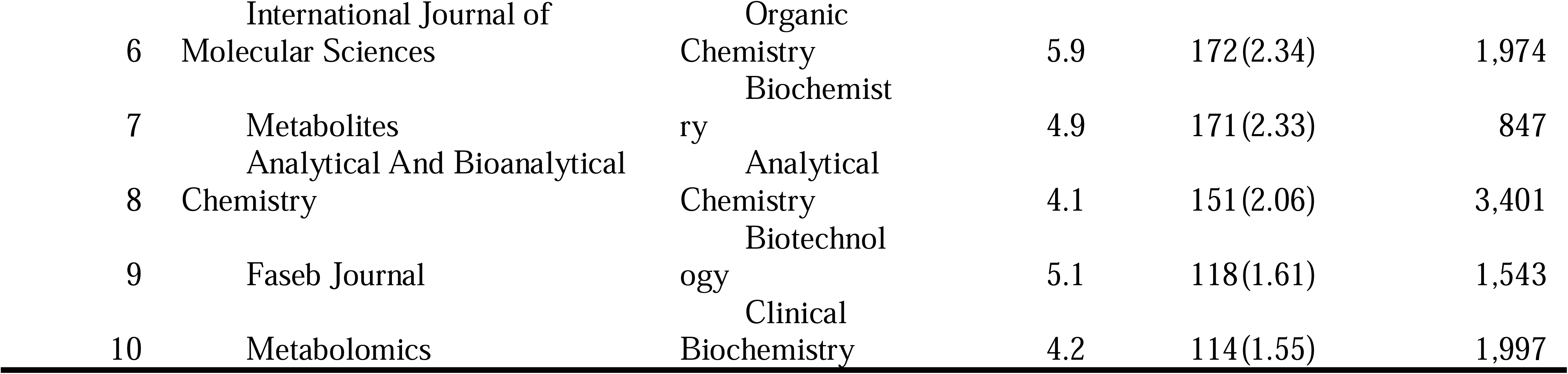
The most productive 10 journals for lipidomics.

A total of 7,338 articles were cited 21,411 times. To better describe the citations of publications, we took the top 15 articles (**Table 4**). The number of citations of the top 15 most cited articles ranged from 149 to 909. “A simple method for the isolation and purification of total lipids from animal tissues”, which was published in *Journal of Biological Chemistry* in 1957, was the most cited article (25,791 citations) (Folch et al., 1957). All of top 15 highest cited articles could be classified into five categories, 1) the extraction method of Lipid (Folch et al., 1957) (Matyash et al., 2008); 2) the lipid identification or quantification (Han and Cheng, 2005) (Cajka and Fiehn, 2014) (Ejsing et al., 2006) (Yang et al., 2009); 3) analytical methods for lipid data (Brügger et al., 1997) (Pluskal et al., 2010) (Tsugawa et al., 2015) (Smith et al., 2006) (Chong et al., 2018); 4) application of lipid database (Sud et al., 2007) (Fahy et al., 2007); and 5) analysis of lipids in plants or yeast (Ejsing et al., 2009) (Welti et al., 2002).

**Table 4.**
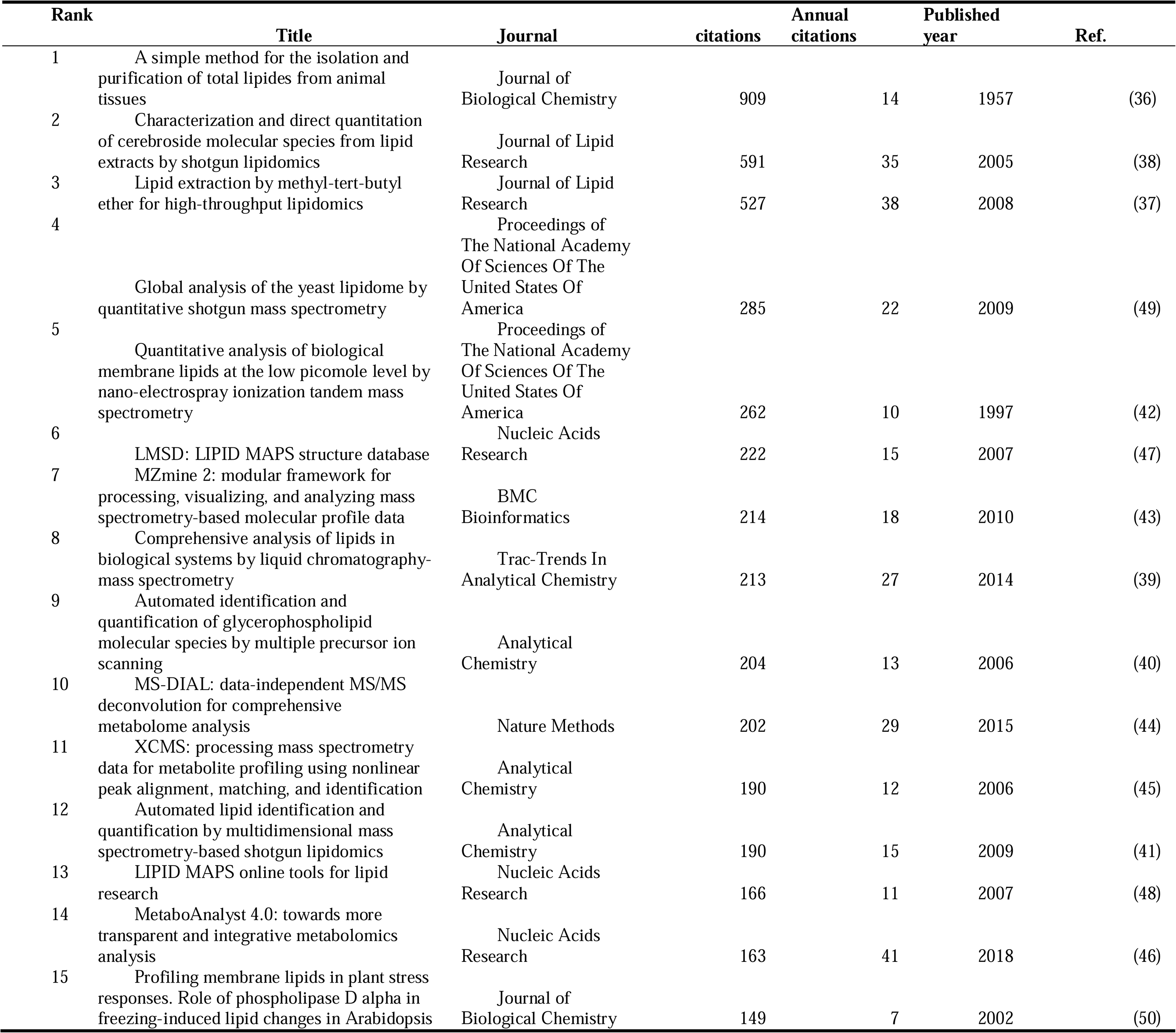
The top 15 highest co-cited references.

### 3.4. Distribution and co-authorship of authors

A total of 45,325 authors with 72,145 appearances published the 7,338 retrieved articles (**Fig. 4**). There were averaged 10 authors in each article. We listed the top 20 most productive authors (**Table 5**). Prof. Xian-Lin Han (146 publications with 7,580 citations) was the most productive, followed by Prof. Oresic Matej (113 publications with 6,791 citations) and Prof. Meikle Peter J (112 publications with 3,892 citations). In the current study, VOSviewer was performed for co-authorship analysis of 1000 authors (each author with more than 5 published articles). The co-authorship network(**Fig. 4**) was divided into 34 clusters represented by different colors. The red cluster was the largest and centered on Prof. Oresic Matej, Ekroos Kim, and Hyotylainen Tuulia. Prof. Xian-Lin Han had the most significant number of cooperating partners (*n* = 38), followed by Prof. Ekroos Kim (*n* = 36), and Liebisch Gerhard (*n* =34).

**Figure 4.**
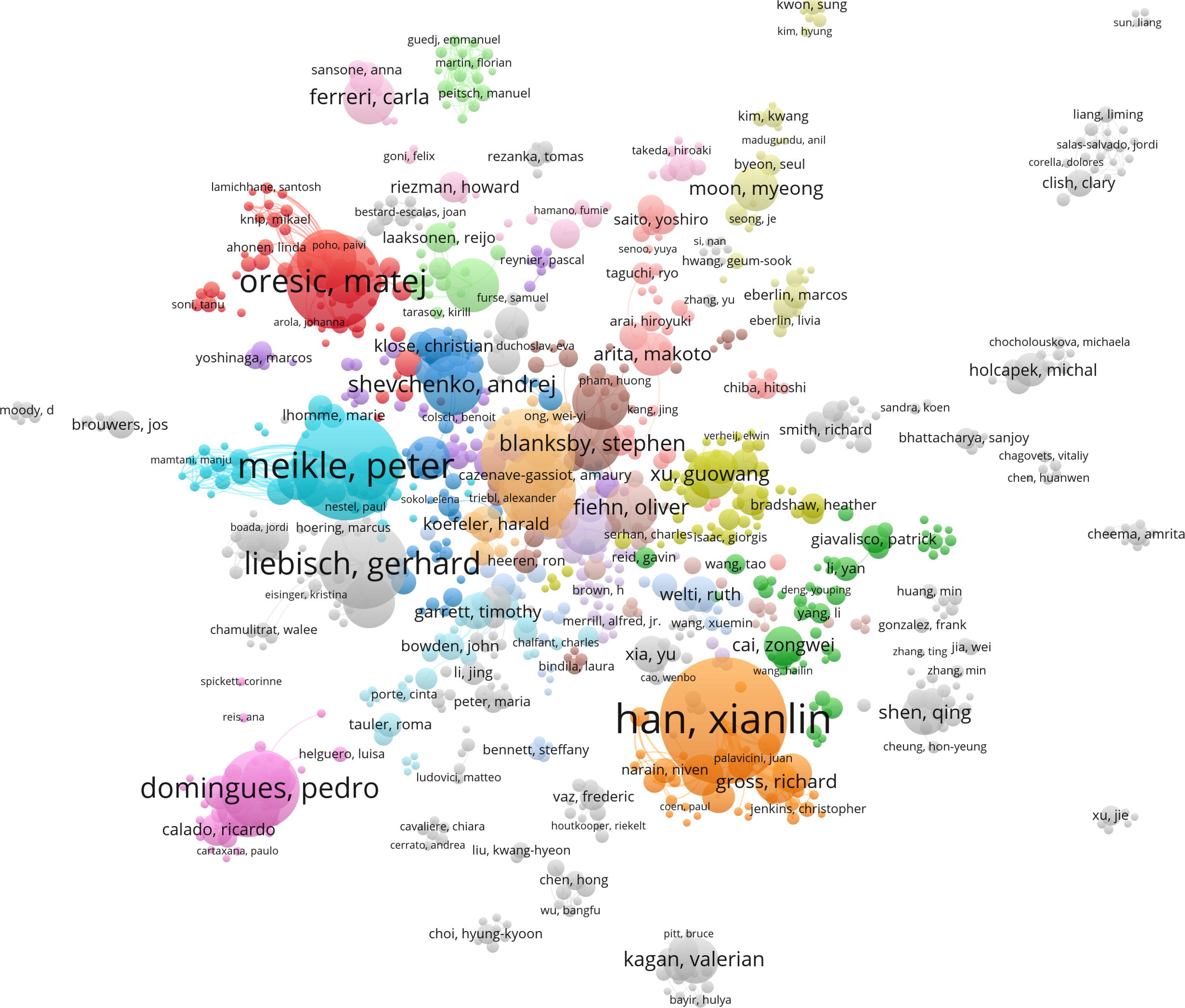
The distributions of co-authors.

**Table 5.**
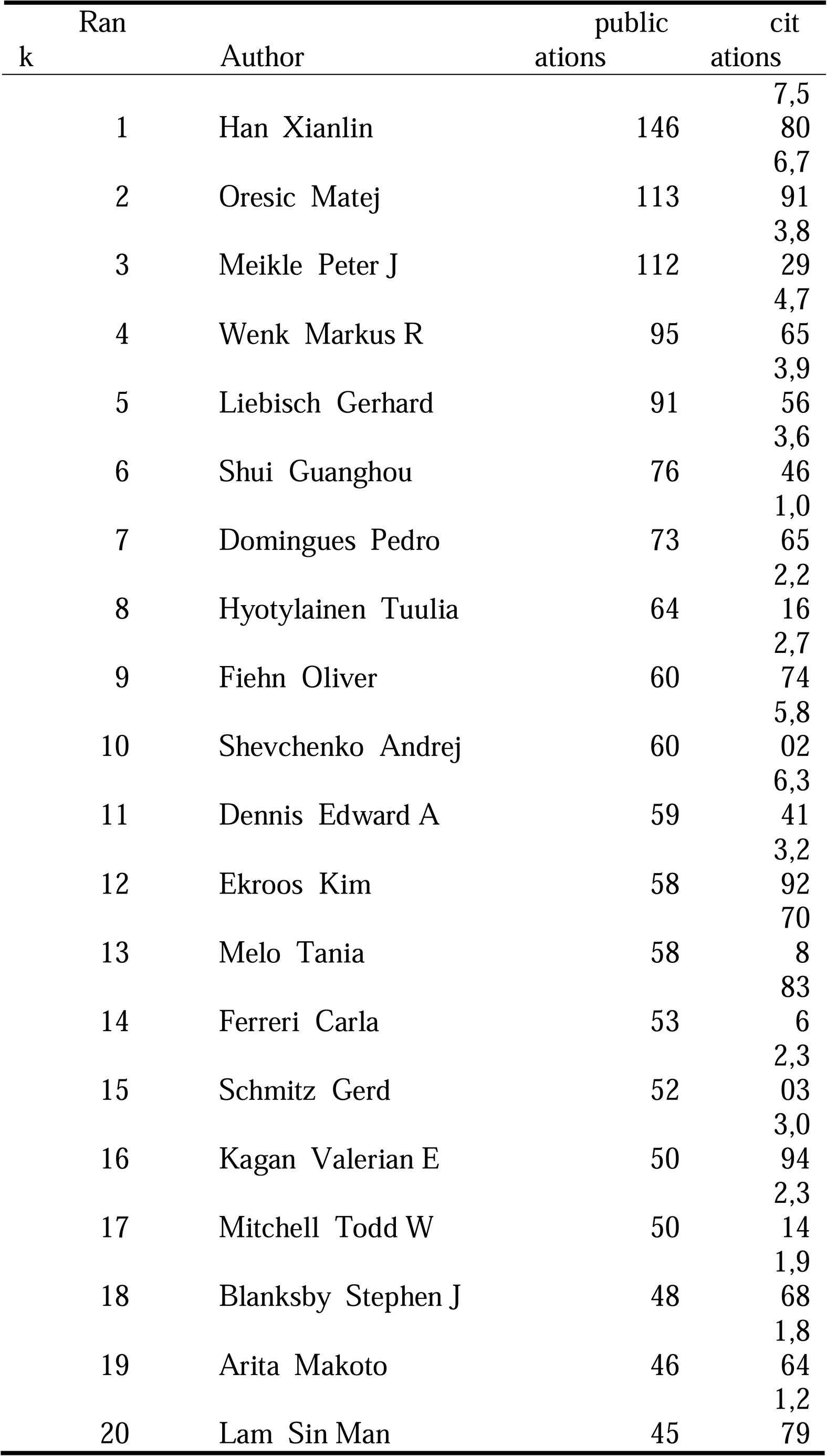
The 20 most productive authors.

### 3.5. Keywords analysis

A total of 878 keywords with 34,650 appearances published the 7,338 retrieved articles and 425 keywords with more than 10 frequencies (**Table S3**). Each article averagely contained 5 keywords. Keywords cover the main topics of publications, and thus high-frequency keywords are well-suited to be selected for co-occurrence analysis (**Fig. 5A**). Furthermore, we caculated the thematic evolution (**Fig. 5B**) of keywords from four period, 2005, 2010, 2015, and 2020, respectively. In the firsted sliced period of 2005, the topic of eicosanoids, electrospray ionization mass spectrometry, sphingolipids, phospholipase a(2), lipidomics, and functional lipidomics turn to mass spectrometry, lipidomics, and inflammation. In the second sliced period of 2010, the topic of apoptosis, ceramide, lipidomics analysis, cardiovascular risk, shotgun lipidomics, lipidomics, metabolomics, urine, lipid proflilling, and eicosanoids turn to phospholipids, lipidomics, inflammation, lipid metabolism. In the third sliced period of 2015, the topic of inflammation, mitochondria, metabolomics, lipidomics, and shotgun lipidomics turn to lipidomics, sphingolipids, lipid metabolism, phospholipids, eicosanoids, and lipidome. In the fourth sliced period of 2020, the topic of inflammation, lipid metabolism, lipidomics, mass spectrometry, and lipidome turn to arachidonic acid, lipidomics, lipidome, metabolomics, lipid metabolism, untargeted lipidomics.

**Fig. 5.**
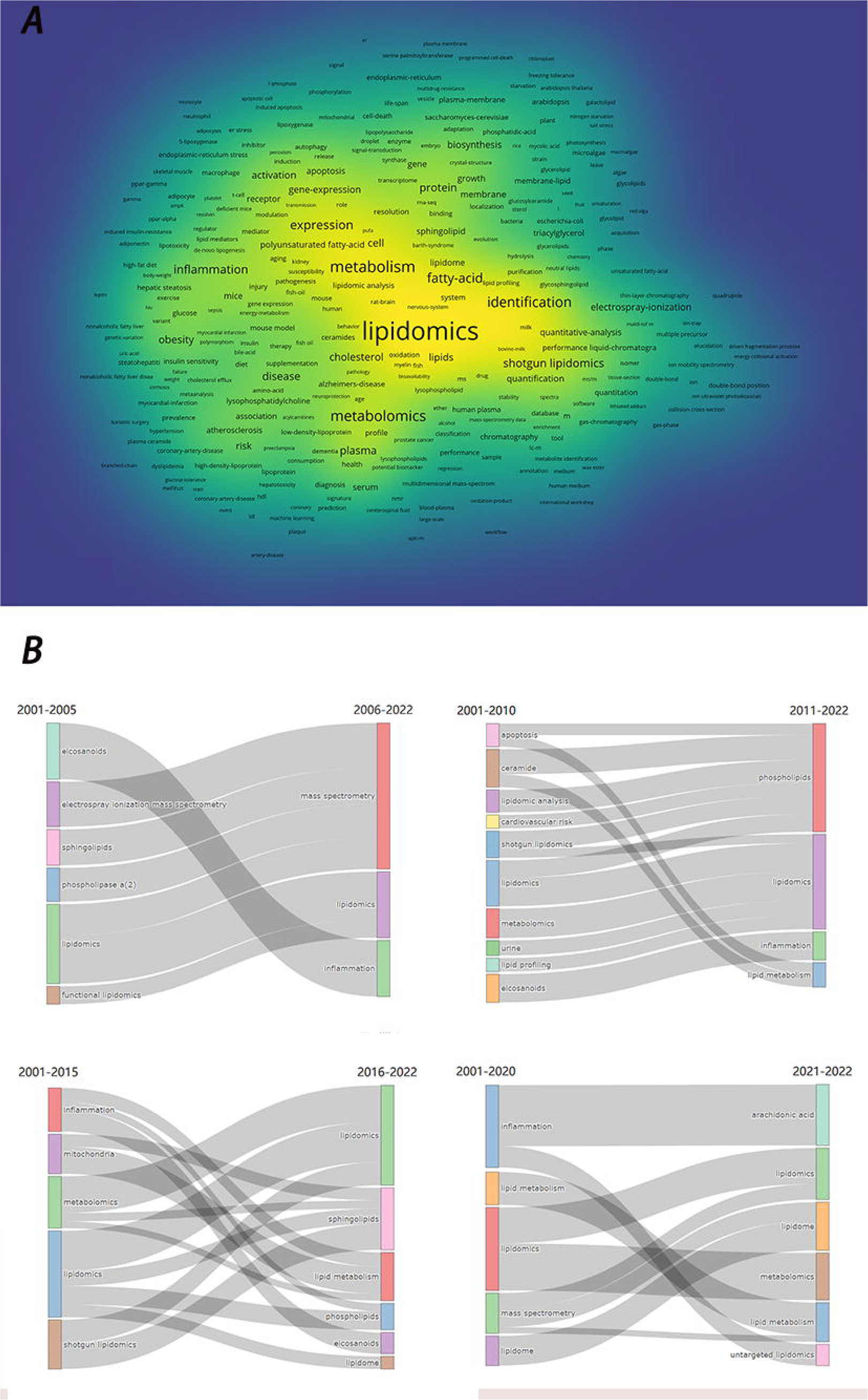
Keywords distributions. A) The distribution of high-frequency keywords. B) The thematic evolution in four periods.

Furthermore, we clustered all of the keywords (**Fig. 6A**) into four types with four colours. The red, green, pink, and blue cluster show the core of shotgun lipidomics, metabolism, identification, and expression, respectively. Following, we clustered four themes (**Fig. 6B**). The niche theme were shotgun lipidomics, tandem mass-spectrometry, and electrospray-ionization. And the motor theme were expression, diseases, and inflammation. The emerging or decling theme were identification, mass-spectrometry, and fatty -acids. Finaly, the basic theme were metabolism, cell, and plasma.

**Fig. 6.**
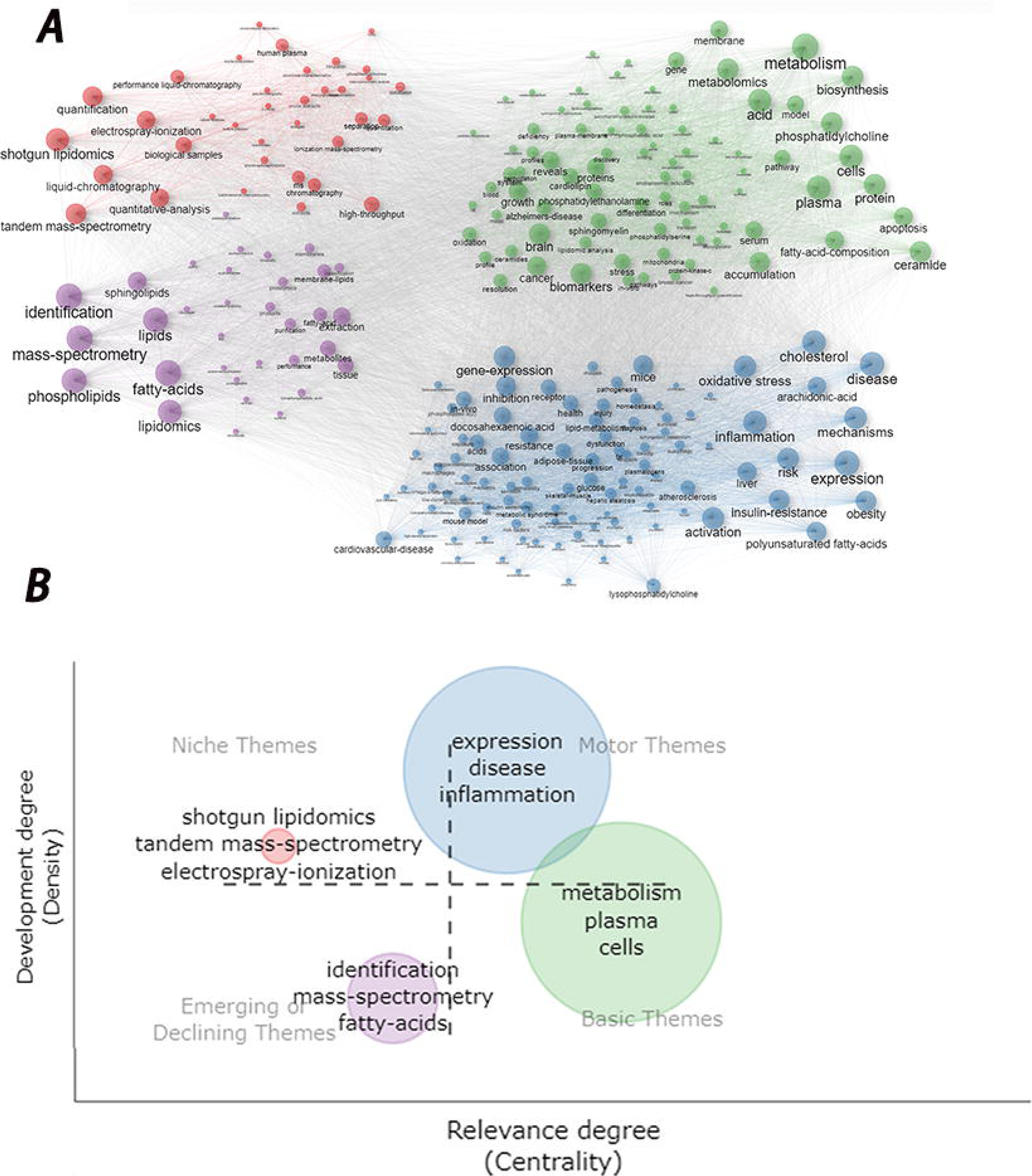
The keywords clusters. A) The keywords clusters in to four color. B) the four theme distribution of keywords.

#### 3.5.1. Lipidomics prefered categories

Although lipidomics was universally classified into eight types, the attention was not received equally. According to LIPID MAPS, we classified the lipidomics-related keywords of minimum 10 counts (**Fig. 7A**) into 5 categories, Fatty Acyls (*n* = 1,830), Glycerophospholipids (*n* = 1,317), Sphingolipids (*n* = 596), Glycerolipids (*n* = 178), and Sterol Lipids(*n* = 342). Fatty Acyls was the most studied batch of lipidomics, followed by Glycerophospholipids. The polyunsaturated fatty (PUFA), arachidonic acid (ARA), and docosahexaenoic acid (DHA) were dominant subsets of Fatty Acyls which indicated a crucial focus in lipidomics. In the studies of Glycerophospholipids, the composed cardinal members were phospholipid, phosphatidylcholine (PC), and phosphatidylethanolamine (PE). The crucial studied members of Sphingolipids, Glycerolipids, and Sterol Lipids were ceramide, triacylglycerol, and cholesterol, respectively.

**Fig. 7.**
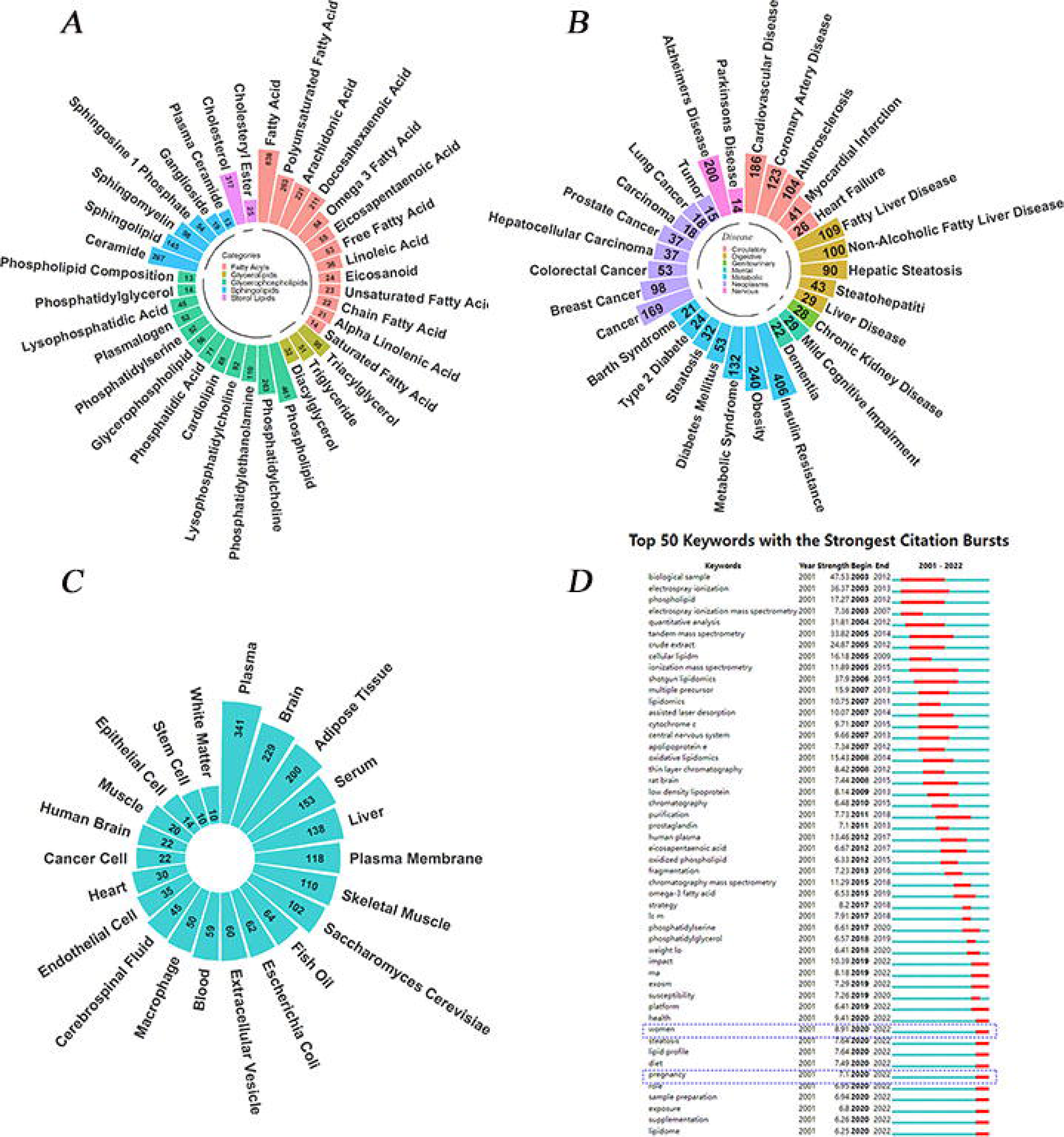
Manual analysis results of keywords frequency. A) The preferred categories of lipidomics. B) The lipidomics-related diseases. C) The lipidomics-related tissues. D) the citation bursts analysis.

#### 3.5.2. Lipidomics-favoured diseases

According to the International Classification of Diseases (ICD, https://icd.who.int/browse11/l-m/en), all of these disease-related keywords were categorized into seven brief types (**Fig. 7B**). We revised several keywords with ICD. For example, we took “insulin-resistance” and “insulin resistance” into a type. And the majority of lipidomics-related diseases were insulin resistance (*n* = 406), followed by obesity (*n* = 240), Alzheimer’s disease (*n* = 200), and cancer (*n* = 169). Metabolic diseases (*n* = 908) were the most frequent lipidomics-related disease, followed by diseases of the circulatory system (*n* = 480).

#### 3.5.3. Lipidomics-favoured tissues

Lipidomics-related researches mainly focus on 22 tissue (**Fig. 7C**). Plasma (*n* = 341) was the most conventional tissue for the extraction of lipid, followed by brain (*n* = 229), adipose tissue (*n* = 200), serum (*n* = 153), and liver (*n* = 138). Plasma was the pervasive tissue for lipidomics extraction, however specific tissue would be utilized in a specific disease. For example, white matter in the brain is lipid-rich tissue which was a target for lipid research (Smith et al., 2022), especially in Alzheimer’s disease (Xu et al., 2020; Zhang et al., 2020a).

#### 3.5.4. Lipidomics-related citation bursts keywords

The citation bursts keywords could predict the potential hot topics in the future with the basement of the current publication. The more duration and more strength in the burst may indicate the powerful prediction of the topic. CiteSpace was performed to filter the burst’s top 50 keywords (**Fig. 7D**), and the longest duration citation burst keywords were “electrospray ionization” and “ionization mass spectrometry”, both with 11 years and strength of 36.37 and 11.89, respectively. Furthermore, the most strength burst keyword was the “biological sample” with a strength of 47.53. In addition, a dotted line box (**Fig. 7A**) embraced two keywords in recent times, “women” and “pregnancy”. Both keywords burst in 2020 with the strength of 8.91 and 7.1, which forecast the potential hot topics in the future.

## 4. Disccusion

### 4.1. Global trends in lipidomics

Since Kishimoto et, al. (Kishimoto et al., 2001) firstly reported lipidomics-related publication in 2001, which developed a nondestructive quantification of neutral lipids with thin-layer chromatography, the lipidomics research had no waves in the following 2 years. Until Han et, al. (Han and Gross, 2003) in 2003 processed ESI mass spectrometry for the bridge of lipidomics, in which lipidomics cached the increasing eyes of scholars. Different from other bibliometrics research (Contreras-Barraza et al., 2021) with acute fluctuations or step-down latency (Xu et al., 2017), the lipidomics-related publications were sustainable surging annually. At present, lipidomics-related research is exponential surging (**Fig. 2**) which indicated more and more (Wenk, 2010) attention was paid. A total of 100 Countries were involved in lipidomics-related publications, and 38 Countries with more than 20 published articles (**Fig. 3A**). The top 3 Countries (USA, China, and Germany) contribute 43.99% of all Countries. China had a late beginning of lipidomics compared with other Countries, but a surprising increase during the 2 decades. Chinese Academy of Sciences was the most productive Country. Among the 382 journals, *Journal of Lipid Research* was the most productive (**Table 3**) and the most highly cited journal. Prof. Xian-Lin Han was the most productive author with 146 publications.

### 4.2. The preferred categories of lipidomics

According to LIPID MAPS, lipidomics was universally classified into 8 categories, but only 5 types (fatty acyls, glycerophospholipids, sphingolipids, glycerolipids, sterol lipids) often catch the studies of publications. Especially Fatty Acyls and Glycerophospholipids seem to get a preference of records (**Fig. 7A)**.

In deeply, the preferred studied subsets of fatty acyls was PUFA. Fatty acids are the most abundant lipid in the body and the major endogenous energy source (Malaisse et al., 1983). The ω-3 and ω-6 PUFA are both major types of PUFA that play an indispensable role in normal human health (Jump, 2002). DHA was a type of PUFA, and PUFA was identified to prevent the incidence of cardiovascular and cerebrovascular diseases (Iso et al., 2001; Strøm et al., 2012). Two system reviews (Enns et al., 2014; Abdelhamid et al., 2018) emphasized that ω-3 PUFA has little or no effect on the arterial disease with cardiovascular events and other serious clinical outcomes. The above research just focused on the PUFA in plasma or serum. Ding D et, al. (Ding et al., 2020) transform the attention to erythrocyte lipid indicated a similar conclusion that the potential cardioprotective roles of very-long-chain PUFA. The moderate intake of more PUFA for individuals means less atherosclerosis risk, but if deficient? Anna LP et, al. (Petursdottir et al., 2008) identified that the absence of ω-3 PUFA is highly associated with memory loss and diminished cognitive function with tryptophan metabolism (Morgese et al., 2020), such as Alzheimer’s disease (Morgese et al., 2020), dementia (Wang et al., 2021), and sleep disorders (Decoeur et al., 2020). The other type of fatty acid studied preferred was ARA which fulfills several physiological functions: compose the phospholipid bilayer of cell membranes, perform a precursor for a crucial category of biologically such as eicosanoids, regulate gene expression, mediate inflammation, and balance vasodilator or vasoconstrictor (Imig, 2020; Wang et al., 2020; Badimon et al., 2021). In a word, Fatty Acyls was the current hot type, especially its subtypes PUFA, DHA, and ARA.

Glycerophospholipids were the second preferred category of lipidomics, especially in PC and PE. Phospholipid, a generic term not a specific type of Glycerophospholipids, compose an important component of the cell membrane and plays an important role in maintaining cell homeostasis. In animals, lecithin or PC is the most abundant, followed by PE (also called cerebral phospholipid). PC, as 80% of major phospholipids, is the surfactant lipid of the lung to maintain alveolar surface tension. The metabolism of PC is associated with cholesterol. Eleni AK et, al (Karavia et al., 2013) indicated that PC/cholesterol acyltransferase activity is an important modulator of processes associated with diet-induced hepatic lipid deposition. PC might decrease inflammation (Machado-Aranda et al., 2013) and regulate immune function. And PC is the major phospholipid comprising 80% of surfactant lipids (Middlekauff et al., 2020) and smoking may down-regulate the content of PC for the progress of chronic obstructive pulmonary disease (Agudelo et al., 2020). PE is the second composition of cell members in humans, enrich in brian or nervous tissue (Karikó et al., 2001).

Above all, fatty acyls and glycerophospholipids are the preferred categories of lipidomics, compared with the other 6 types, which indicated a current trend, especially in the subsets of PUFA, ARA, DHA, PC, and PE. Furthermore, ceramide and cholesterol are also popular in research (**Fig. 7A**).

### 4.3. The lipidomics-favoured diseases

According to the ICD, the keywords of more than 10 frequencies were summarized and the lipidomics-related diseases were categorized into 7 types (**Fig. 7B**). Insulin resistance, obesity, and Alzheimer’s disease were the most representative of endocrine, nutritional or metabolic diseases, diseases of the circulatory system, and diseases of the nervous system.

Insulin resistance is a typical clinical manifestation and one of the criteria for diagnoses of type 2 diabetes (Mojiminiyi et al., 2007) that most research focuses on both diseases without discrimination. The insulin resistance-lipidome alterations had been identified as the characterizing factors of non-alcoholic steatohepatitis (Guerra et al., 2022). Insulin resistance is associated with obesity. Chloe W et, al. (Wilkin et al., 2021) focused on the phospholipidome of the mononuclear cells in both disease and phospholipidome remodeling insulin resistance could disrupt the cell membranes and immune dysfunction. In type 2 diabetes, RBC membrane lipidome with increased cholesterol, total sphingolipids, sphingomyelin, and glycolipids, but decreased phospholipids may trigger impairment of membrane fluidity and rigidity (Kostara et al., 2021). Altered lipidomics in type 2 diabetes is comprehensive rather than single type (higher triacylglycerols and diacyl-phospholipids but lower alkyl acyl phosphatidylcholines) may be conducive to the forecast of the progress of type 2 diabetes (Suvitaival et al., 2018). Interestingly, the altered lipidomics in insulin resistance may be associated with sex (Beyene et al., 2020a). In a word, lipidomics can help diagnose insulin resistance/type 2 diabetes and predict disease progression, the timely intervention of lipidomics might reverse the disease (Zobel et al., 2021) which might trigger the preference for research.

Previous research had reported obesity had potential progress of insulin resistance, type 2 diabetes, and even pancreatic cancer (Bao et al., 2011). Obese children had significantly altered phospholipidome compared with normal-weight children (Anjos et al., 2019). The altered lipidomics may inherit by offspring (León-Aguilar et al., 2019), obese mothers exhibited a significant reduction in the total abundance of ceramides (Cer) in plasma, mainly of Cer (d18:1/20:0), Cer (d18:1/22:0), Cer (d18:1/23:0), and Cer (d18:1/24:0) and the altered ceramides might deliver to offspring even with a normal diet. Similar to insulin resistance, Habtamu BB et, al. (Beyene et al., 2020b) emphasize that obesity altered lipidomics was associated with sex and age. Specific classes of ether-phospholipids and lysophospholipids (calculated as the sum composition of the species within the class) were inversely associated with age in men only (Beyene et al., 2020b). The obese postmenopausal women had higher triacylglycerol and lower lysoalkyl phosphatidylcholine species compared with premenopausal women (Beyene et al., 2020b). In the erythrocyte membrane, the higher interfacial fluidity in obese patient erythrocyte might result from the switch from ω-3 to ω-6 lipids (Sot et al., 2022). In a word, obesity maintains a status of altered lipidomics, the current research focused on both the manifestation in macroscopic phenomena (such as sex and age) and microcosmic characteristics such as erythrocyte membrane, the transformation of lipidomics with medicine (Fernández-Arroyo et al., 2019) may regulate cardiovascular disease risk.

Alzheimer’s disease was the third prefereed disease for lipidomics. The target for Alzheimer’s disease was brian which enriches the lipids, especially in PE and maybe this is the reason for the popularity of lipidomics research in this disease. The plasma lipids (sphingomyelins, cholesterol esters, PC, PE, phosphatidylinositols, and triglycerides) are dysregulated in Alzheimer’s disease patients which may help discriminate them from healthy controls (Liu et al., 2021). The association of lipidomics signatures in blood with clinical progression related to preclinical and prodromal Alzheimer’s Disease (Sakr et al., 2022). The intervention of Rhodiola crenulate in Alzheimer’s Disease rats can remodel the distribution of lipidomics and protect the Alzheimer’s Disease rats (Sun et al., 2020).

To sum up, insulin resistance, obesity, and Alzheimer’s disease were the preferred disease of lipidomics research, and previous studies attributed to exploring the diagnosis, forecast progression, and intervention to better know these diseases. Other diseases in our research such as cancer, and cardiovascular disease et, al were also studied extensively.

### 4.4. The lipidomics-favoured tissue

Lipidomics-related researches mainly focus on 22 tissue (**Fig. 7C**). The plasma was the top 1 tissue, followed by the brain, adipose tissue, serum et, al. Diverse tissue was performed for different diseases or models. Plasma was widely applied for clinical trials or animal experiments such as diabetes (Forouhi et al., 2014), coronary heart disease (Si et al., 2021), pneumonia (Snider et al., 2021), and so on. Interestingly, although serum and plasma were both derived from blood and serum has both a convenient handle method and relatively high concentrations of lipidomics (Rajan et al., 2019), the extraction of lipid was preferred to plasma. This phenomenon might be attributed to serum having relatively low levels of the same amount of blood. Brain-lipidomics were always specific diseases, such as alcoholic brain injury (Smith et al., 2022), Alzheimer’s disease (Lauer et al., 2021), and neuro inflammatory disease (Derada Troletti et al., 2021). Adipose tissue contains abundant lipids and can be classified into two subtypes, white and brown adipose tissue (Grzybek et al., 2019). Previous research focused on both types of adipose tissue and compared these differences (Fuse et al., 2020; Lange et al., 2021). In a word, although plasma was the majority selection for lipidomics extraction, other types of tissue may be specific for models/diseases such as the brain for Alzheimer’s disease, liver for nonalcoholic hepatitis (Bissig-Choisat et al., 2021), exercise for skeletal muscle (Mitchell et al., 2010), and so on.

### 4.5. The potential hotspots for the future lipidomics

Among 50 lipidomics-related citation bursts, keywords (**Fig. 7D**) show the longest duration citation burst keywords were “electrospray ionization” and “ionization mass spectrometry”. Furthermore, the most strength burst keyword was the “biological sample” which emphasizes the crucial choice of the sample. In addition, a dotted line box (**Figure 7d**) embraced two keywords in recent times, “women” and “pregnancy” respectively.

Several studies reported specific lipidomics for women. For example, in insulin resistance, the alkylphosphatidylcholine (10), alkenylphosphatidylcholine (23), and alkylphosphatidylethanolamine (6) were associated with men only, but phosphatidylcholine (7) and sphingomyelin (5) were associated in women only(Beyene et al., 2020a). The obese postmenopausal women had higher triacylglycerol and lower lysoalkyl phosphatidylcholine species compared with premenopausal women (Beyene et al., 2020b). Although these studies reported these phenomena but without lucubration which indicated women may have different lipidomics profiles compared to men. So women have characteristic lipidomics profiles and maybe a potential hotspot in the future.

Pregnancy is a special period that may incidence specific diseases such as gestational diabetes, gestational hypertension, postpartum depression, and so on. For gestational diabetes, Samuel Furse et, al. (Furse et al., 2021) reported lipid metabolism modulated in a healthy pregnancy, and the timing of these changes is altered in gestational diabetes pregnancies. Both postpartum depression and dementia are prevalent mental disorders, ω −3 PUFA can protect against both diseases (De Vriese et al., 2003; Lim et al., 2006). In dementia, the medical treatment (Sun et al., 2021) may be related to the lipidomics profile. To our knowledge, postpartum depression has no such study. In major antenatal depression, the cholesterol sulfate and PC (18:2 (2E, 4E)/0:0) may be effective and specific lipidic biomarkers for the prediction of antenatal depression (Wu et al., 2019). The above studies for pregnancy just for better biomarkers in lipidomics but no intervention of lipidomics which indicated that pregnancy-related lipidomics has a giant potential hotspot in the future.

Bibliometric analysis was performed on the evolution and trends of lipidomics. Though relatively comprehensive and objective the bibliometric analysis was, some inevitable limitations similar to previous studies (Briganti et al., 2019; Zyoud, 2019; Peng et al., 2022). Lipidomics-related publications in this study were only downloaded from the WOScc database. Even the most commonly used and authoritative comprehensive database the WOSCC is (Boudry et al., 2018; Zhang et al., 2020b), minority publications without being included. Another limitation ignore the quality of publications and we took high or low-quality publications in the same weight. Regarding the authors, the name of the authors was automatically extracted from CiteSpace and VOSviewer which may not be extracted correctly. If authors use different name spellings or multiple names, the extraction of names will be inaccurate.

## 5. Conclussions

To the best of our knowledge, this work provided comprehensive analysis of lipidomics from a bibliometric analysis perspective for the first time. Our bibliometrics portrayed a visualization of lipidomics by reviewing publication records over the past two decades. This research summarizes the temporal trends and evolution, global collaboration patterns, and potential hotspots in the future. These findings enable the research community to identify the emerging themes and frontiers which can guide future research on lipidomics.

## Data Availability

All data produced in the present study are available upon reasonable request to the authors.

## Abbreviations

PUFA: Polyunsaturated fatty
ARA: Arachidonic acid
DHA: Docosahexaenoic acid
ICD: International Classification of Diseases
PC: Phosphatidylcholine
PE: Phosphatidylethanolamine

## Ethics approval and consent to participate

Not applicable.

## Consent for publication

Not applicable.

