## Supplementary material for "Bibliometric and visualized mapping: two decades of lipidomics, with special focus on pregancy and women": Table S1, Table S2, Table S3.

**Table S1.** The national distribution of lipidomics-related articles

**Table S2.** The 234 institutions published more than 10 lipidomics-related records

**Table S3.** The 425 keywords with more than 10 frequencies

**Table S1**. The national distribution of lipidomics-related articles

| Rank | | Publications(Percentage%) | Beginning time | Country |
| --- | --- | --- | --- | --- |
| 1 | 2577(22.73) | | 2002 | USA |
| 2 | 1517(13.38) | | 2005 | China |
| 3 | 893(7.88) | | 2006 | Germany |
| 4 | 574(5.06) | | 2002 | England |
| 5 | 441(3.89) | | 2003 | France |
| 6 | 409(3.61) | | 2005 | Italy |
| 7 | 399(3.52) | | 2006 | Australia |
| 8 | 394(3.48) | | 2007 | Spain |
| 9 | 358(3.16) | | 2001 | Japan |
| 10 | 324(2.86) | | 2004 | Canada |
| 11 | 295(2.6) | | 2005 | Netherlands |
| 12 | 247(2.18) | | 2005 | Finland |
| 13 | 235(2.07) | | 2002 | Sweden |
| 14 | 201(1.77) | | 2006 | Switzerland |
| 15 | 178(1.57) | | 2006 | South Korea |
| 16 | 173(1.53) | | 2006 | Singapore |
| 17 | 170(1.5) | | 2005 | Austria |
| 18 | 169(1.49) | | 2005 | Denmark |
| 19 | 163(1.44) | | 2008 | Brazil |
| 20 | 141(1.24) | | 2010 | Poland |
| 21 | 137(1.21) | | 2011 | Portugal |
| 22 | 126(1.11) | | 2006 | Czech Republic |
| 23 | 103(0.91) | | 2006 | Belgium |
| 24 | 101(0.89) | | 2005 | Russia |
| 25 | 90(0.79) | | 2007 | India |
| 26 | 74(0.65) | | 2007 | Scotland |
| 27 | 74(0.65) | | 2009 | Norway |
| 28 | 63(0.56) | | 2007 | Wales |
| 29 | 58(0.51) | | 2010 | Israel |
| 30 | 48(0.42) | | 2008 | Greece |
| 31 | 45(0.4) | | 2014 | Saudi Arabia |
| 32 | 43(0.38) | | 2010 | Hungary |
| 33 | 43(0.38) | | 2007 | Ireland |
| 34 | 35(0.31) | | 2014 | Egypt |
| 35 | 31(0.27) | | 2008 | New Zealand |
| 36 | 29(0.26) | | 2011 | Argentina |
| 37 | 21(0.19) | | 2013 | Mexico |
| 38 | 20(0.18) | | 2014 | Turkey |
| 39 | 19(0.17) | | 2008 | Croatia |
| 40 | 18(0.16) | | 2016 | North Ireland |
| 41 | 17(0.15) | | 2007 | South Africa |
| 42 | 16(0.14) | | 2013 | Thailand |
| 43 | 16(0.14) | | 2017 | Vietnam |
| 44 | 16(0.14) | | 2018 | Colombia |
| 45 | 15(0.13) | | 2015 | Pakistan |
| 46 | 14(0.12) | | 2012 | Chile |
| 47 | 12(0.11) | | 2009 | Malaysia |
| 48 | 11(0.1) | | 2012 | Iran |
| 49 | 11(0.1) | | 2015 | Iceland |
| 50 | 11(0.1) | | 2009 | Slovenia |
| 51 | 11(0.1) | | 2013 | Qatar |
| 52 | 10(0.09) | | 2009 | Serbia |
| 53 | 10(0.09) | | 2014 | Bulgaria |
| 54 | 9(0.08) | | 2016 | Bangladesh |
| 55 | 9(0.08) | | 2015 | Lebanon |
| 56 | 8(0.07) | | 2018 | Romania |
| 57 | 7(0.06) | | 2010 | Luxembourg |
| 58 | 7(0.06) | | 2017 | Indonesia |
| 59 | 7(0.06) | | 2013 | Slovakia |
| 60 | 6(0.05) | | 2008 | Cyprus |
| 61 | 5(0.04) | | 2015 | Belarus |
| 62 | 5(0.04) | | 2016 | Tunisia |
| 63 | 5(0.04) | | 2014 | Ethiopia |
| 64 | 4(0.04) | | 2015 | U Arab Emirates |
| 65 | 4(0.04) | | 2018 | Estonia |
| 66 | 3(0.03) | | 2018 | Ecuador |
| 67 | 3(0.03) | | 2020 | Nigeria |
| 68 | 3(0.03) | | 2020 | Costa Rica |
| 69 | 3(0.03) | | 2017 | Uganda |
| 70 | 2(0.02) | | 2016 | Nepal |
| 71 | 2(0.02) | | 2016 | Algeria |
| 72 | 2(0.02) | | 2020 | Mali |
| 73 | 2(0.02) | | 2021 | Ghana |
| 74 | 2(0.02) | | 2019 | Morocco |
| 75 | 2(0.02) | | 2019 | Bahrain |
| 76 | 2(0.02) | | 2015 | Georgia |
| 77 | 2(0.02) | | 2019 | Gambia |
| 78 | 2(0.02) | | 2018 | Uruguay |
| 79 | 2(0.02) | | 2019 | Venezuela |
| 80 | 2(0.02) | | 2017 | Rwanda |
| 81 | 1(0.01) | | 2020 | Myanmar |
| 82 | 1(0.01) | | 2019 | St Kitts & Nevi |
| 83 | 1(0.01) | | 2020 | Yemen |
| 84 | 1(0.01) | | 2018 | Madagascar |
| 85 | 1(0.01) | | 2018 | Iraq |
| 86 | 1(0.01) | | 2020 | French Guiana |
| 87 | 1(0.01) | | 2007 | Senegal |
| 88 | 1(0.01) | | 2019 | Ukraine |
| 89 | 1(0.01) | | 2021 | Jordan |
| 90 | 1(0.01) | | 2021 | Latvia |
| 91 | 1(0.01) | | 2019 | Lithuania |
| 92 | 1(0.01) | | 2021 | Sudan |
| 93 | 1(0.01) | | 2019 | Sierra Leone |
| 94 | 1(0.01) | | 2021 | Kenya |
| 95 | 1(0.01) | | 2007 | Burkina Faso |
| 96 | 1(0.01) | | 2017 | Syria |
| 97 | 1(0.01) | | 2020 | Gabon |
| 98 | 1(0.01) | | 2020 | Philippines |
| 99 | 1(0.01) | | 2021 | Monaco |
| 100 | 1(0.01) | | 2021 | Montenegro |
| 101 | 1(0.01) | | 2019 | Tanzania |

**Table S2**. The 234 institutions published more than 10 lipidomics-related records

| rank | Institution | Country | Publications | Beginning time |
| --- | --- | --- | --- | --- |
| 1 | Chinese Academy of Sciences | China | 321 | 2007 |
| 2 | University of Washington | USA | 169 | 2003 |
| 3 | National University of Singapore | Singapore | 128 | 2006 |
| 4 | University of Helsinki | Finland | 113 | 2005 |
| 5 | University of Melbourne | Australia | 106 | 2006 |
| 6 | Baker Heart and Diabetes Institute | Australia | 105 | 2017 |
| 7 | University of California San Diego | USA | 102 | 2005 |
| 8 | University of Aveiro | Portugal | 100 | 2012 |
| 9 | Harvard University | USA | 100 | 2002 |
| 10 | University of California, Davis | USA | 97 | 2011 |
| 11 | Harvard Medical School | USA | 94 | 2016 |
| 12 | Consiglio Nazionale delle Ricerche | Italy | 83 | 2005 |
| 13 | University of Cambridge | England | 82 | 2007 |
| 14 | Peking University | China | 78 | 2012 |
| 15 | University of Florida | USA | 77 | 2017 |
| 16 | University of Pittsburgh | USA | 71 | 2007 |
| 17 | Purdue University | USA | 71 | 2010 |
| 18 | Leiden university | Netherlands | 71 | 2006 |
| 19 | Karolinska Inst Stockholm Sweden | Sweden | 71 | 2011 |
| 20 | University of Turku | Finland | 70 | 2011 |
| 21 | Monash University | Australia | 68 | 2017 |
| 22 | University of Tokyo | Japan | 67 | 2005 |
| 23 | Vanderbilt University | USA | 62 | 2003 |
| 24 | University of Sao Paulo | Brazil | 60 | 2012 |
| 25 | University of Michigan | USA | 59 | 2016 |
| 26 | University of Copenhagen | Denmark | 59 | 2015 |
| 27 | Imperial College London | England | 58 | 2016 |
| 28 | Chinese Academy of Agricultural Sciences | China | 57 | 2015 |
| 29 | Duke University | USA | 56 | 2005 |
| 30 | University of Regensburg | Germany | 55 | 2006 |
| 31 | King's College London | England | 54 | 2013 |
| 32 | Consejo Superior de Investigaciones Cientificas | Spain | 51 | 2012 |
| 33 | Max Planck Institute of Molecular Cell Biology and Genetics | Germany | 53 | 2009 |
| 34 | Columbia University | USA | 53 | 2007 |
| 35 | VTT Technical Research Centre of Finland | Finland | 52 | 2007 |
| 36 | University of Oxford | England | 51 | 2004 |
| 37 | Brigham and Women’s Hospital | USA | 51 | 2002 |
| 38 | Tsinghua University | China | 48 | 2015 |
| 39 | Fudan University | China | 48 | 2014 |
| 40 | University of Pennsylvania | USA | 47 | 2003 |
| 41 | University of California, Los Angeles | USA | 46 | 2011 |
| 42 | Shanghai Jiao Tong University | China | 46 | 2014 |
| 43 | Zhejiang University | China | 44 | 2005 |
| 44 | Johns Hopkins University | USA | 44 | 2013 |
| 45 | University of Tübingen | Germany | 42 | 2007 |
| 46 | University of Wollongong | Australia | 42 | 2007 |
| 47 | University of Amsterdam | Netherlands | 42 | 2011 |
| 48 | Technical University of Munich | Germany | 41 | 2017 |
| 49 | Ohio State University | USA | 40 | 2007 |
| 50 | National Institute for Health and Medical Research | France | 40 | 2008 |
| 51 | University of Barcelona | Spain | 39 | 2012 |
| 52 | Hong Kong Baptist University | China | 39 | 2014 |
| 53 | Georgetown University | USA | 39 | 2012 |
| 54 | Waters Corporation | USA | 38 | 2014 |
| 55 | University of California,Irvine | USA | 38 | 2005 |
| 56 | Seoul National University | South Korea | 38 | 2016 |
| 57 | Keio University | Japan | 38 | 2012 |
| 58 | Cardiff University | England | 38 | 2007 |
| 59 | Medical University of Graz | Austria | 37 | 2011 |
| 60 | Instituto de Salud Carlos III, Madrid | Spain | 37 | 2015 |
| 61 | Charles University, Prague | Czech Republic | 37 | 2014 |
| 62 | University of New South Wales | Australia | 36 | 2019 |
| 63 | University of Campinas | Brazil | 36 | 2014 |
| 64 | ABO Akademi University | Finland | 36 | 2006 |
| 65 | Yonsei University | South Korea | 35 | 2009 |
| 66 | University of Montpellier | France | 35 | 2017 |
| 67 | Max Planck Institute of Molecular Plant Physiology | Germany | 35 | 2011 |
| 68 | University of Groningen | Netherlands | 34 | 2014 |
| 69 | University of Birmingham | England | 34 | 2005 |
| 70 | Orebro University | Sweden | 34 | 2017 |
| 71 | Harvard T.H.Chan School of Public Health | USA | 34 | 2016 |
| 72 | Capital Medical University | China | 34 | 2014 |
| 73 | Virginia Commonwealth University | USA | 33 | 2010 |
| 74 | Utrecht University | Netherlands | 33 | 2005 |
| 75 | University of Sydney | Australia | 33 | 2010 |
| 76 | University of Colorado Denver | USA | 33 | 2010 |
| 77 | Chinese Academy of Medical Sciences | China | 32 | 2013 |
| 78 | Dresden University of Technology | Germany | 31 | 2011 |
| 79 | Pacific Northwest National Laboratory | USA | 30 | 2016 |
| 80 | Maastricht University | Netherlands | 30 | 2018 |
| 81 | Lipotype Gesellschaft mit beschrankter Haftung | Germany | 30 | 2016 |
| 82 | Centre national de la recherche scientifique | France | 30 | 2003 |
| 83 | University of Toronto | Canada | 29 | 2016 |
| 84 | University of North Carolina | USA | 29 | 2004 |
| 85 | University Leipzig | Germany | 29 | 2006 |
| 86 | Sanford-Burnham Medical Research Institute | USA | 29 | 2011 |
| 87 | Kansas State University | USA | 29 | 2006 |
| 88 | Hokkaido University | Japan | 29 | 2005 |
| 89 | University of Colorado Boulder | USA | 28 | 2005 |
| 90 | University of Southern Denmark | Denmark | 28 | 2013 |
| 91 | University of Milan | Italy | 28 | 2011 |
| 92 | University of Manchester | England | 28 | 2002 |
| 93 | Indiana University | USA | 28 | 2005 |
| 94 | Helmholtz Zentrum Munchen | Germany | 27 | 2014 |
| 95 | Czech Academy of Sciences | Czech Republic | 27 | 2018 |
| 96 | University of Munich | Germany | 26 | 2016 |
| 97 | University of Wisconsin | USA | 26 | 2018 |
| 98 | Sun Yat-sen University | China | 26 | 2019 |
| 99 | Stanford University | USA | 26 | 2019 |
| 100 | Queensland University of Technology | Australia | 26 | 2015 |
| 101 | University of Gothenburg | Sweden | 25 | 2010 |
| 102 | Polish Academy of Sciences | Poland | 25 | 2016 |
| 103 | Georgia Institute of Technology | USA | 25 | 2005 |
| 104 | University of Southampton | England | 24 | 2004 |
| 105 | Russian Academy of Science | Russia | 24 | 2017 |
| 106 | Pierre and Marie Curie University | France | 24 | 2009 |
| 107 | Osaka University | Japan | 24 | 2012 |
| 108 | Nanjing university of Chinese medicine | China | 24 | 2019 |
| 109 | Massachusetts General Hospital | USA | 24 | 2011 |
| 110 | Zhejiang Gongshang University | China | 23 | 2020 |
| 111 | University of Texas Health Science Center at San Antonio | USA | 23 | 2018 |
| 112 | University of Queensland | Australia | 23 | 2015 |
| 113 | University of Maryland | USA | 23 | 2014 |
| 114 | University of Geneva | Switzerland | 23 | 2011 |
| 115 | Wenzhou Medical University | China | 22 | 2020 |
| 116 | University of Alabama at Birmingham | USA | 22 | 2018 |
| 117 | McGill Universit | Canada | 22 | 2013 |
| 118 | King Abdulaziz University | Saudi Arabia | 22 | 2015 |
| 119 | Humboldt University of Berlin | Germany | 22 | 2007 |
| 120 | Paris-Saclay University | France | 21 | 2017 |
| 121 | Heidelberg University | Germany | 21 | 2012 |
| 122 | University of hong kong | China | 20 | 2015 |
| 123 | Tampere University Hospital | Finland | 20 | 2006 |
| 124 | National Institute of Health Sciences | USA | 20 | 2013 |
| 125 | Mayo Clin | USA | 20 | 2015 |
| 126 | Icahn School of Medicine at Mount Sinai | USA | 20 | 2016 |
| 127 | Deakin University | Australia | 20 | 2013 |
| 128 | Cornell University | USA | 20 | 2016 |
| 129 | Zora Biosciences Oy | Finland | 19 | 2013 |
| 130 | University of Tampere | Finland | 19 | 2013 |
| 131 | Thermo Fisher Scientific | USA | 19 | 2013 |
| 132 | SichuanUniversity | China | 19 | 2020 |
| 133 | Ocean University of China | China | 19 | 2020 |
| 134 | Michigan State University | USA | 19 | 2009 |
| 135 | Huazhong University of Science and Technology | China | 19 | 2019 |
| 136 | University of Texas Southwestern Medical Center | USA | 18 | 2017 |
| 137 | University of Georgia | USA | 18 | 2005 |
| 138 | Steno Diabetes Center Copenhagen | Denmark | 18 | 2018 |
| 139 | Sorbonne University | France | 18 | 2019 |
| 140 | Shandong University | China | 18 | 2013 |
| 141 | Sanford Burnham Prebys Medical Discovery Institute | USA | 18 | 2016 |
| 142 | Rovira i Virgili University | spain | 18 | 2015 |
| 143 | Northwestern University | China | 18 | 2010 |
| 144 | National Institute for Health and Welfare | Finland | 18 | 2010 |
| 145 | Jinan University | China | 18 | 2019 |
| 146 | Graz University of Technology | Austria | 18 | 2008 |
| 147 | Ecole Polytechnique Federale de Lausanne | Switzerland | 18 | 2014 |
| 148 | Wayne State University | USA | 17 | 2010 |
| 149 | University of Pardubice | Czech Republic | 17 | 2012 |
| 150 | Peking Union Medical College Hospital | China | 17 | 2014 |
| 151 | University of Western Australia | Australia | 16 | 2016 |
| 152 | University of Toulouse | France | 16 | 2018 |
| 153 | University of Lausanne | Switzerland | 16 | 2010 |
| 154 | University of Bologna | Italy | 16 | 2019 |
| 155 | National Institute for Agricultural Research | Morocco | 16 | 2008 |
| 156 | National Cancer Institute | USA | 16 | 2012 |
| 157 | University of Arizona | USA | 15 | 2005 |
| 158 | University of Miami | USA | 15 | 2013 |
| 159 | University of Cologne | Germany | 15 | 2016 |
| 160 | University of California, San Francisco | USA | 15 | 2011 |
| 161 | University of California, Berkeley | USA | 15 | 2005 |
| 162 | University of British Columbia | Canada | 15 | 2020 |
| 163 | University of Bordeaux | France | 15 | 2012 |
| 164 | University of Adelaide | Australia | 15 | 2013 |
| 165 | New York University | USA | 15 | 2017 |
| 166 | Medical University of Vienna | Austria | 15 | 2012 |
| 167 | KyungHee University | Republic of Korea | 15 | 2016 |
| 168 | Helsinki University Hospital | Finland | 15 | 2012 |
| 169 | German Centre for Diabetes Research | Germany | 15 | 2014 |
| 170 | Central South University | China | 15 | 2020 |
| 171 | apan Science and Technology Agency | Japan | 15 | 2008 |
| 172 | University of the Basque Country | Spain | 14 | 2011 |
| 173 | University of Kentucky | USA | 14 | 2011 |
| 174 | Stony Brook University-SUNY | USA | 14 | 2016 |
| 175 | RIkagaku KENkyusho/Institute of Physical and Chemical Research | Japan | 14 | 2016 |
| 176 | North Carolina State University | USA | 14 | 2020 |
| 177 | National Taiwan University | China | 14 | 2014 |
| 178 | Medical University of South Carolina | USA | 14 | 2019 |
| 179 | Broad Institute of MIT and Harvard | USA | 14 | 2019 |
| 180 | AstraZeneca | England | 14 | 2012 |
| 181 | University of Ottawa | Canada | 13 | 2013 |
| 182 | University Bonn | Germany | 13 | 2006 |
| 183 | Paris Descartes University | France | 13 | 2009 |
| 184 | Nanjing Medical University | China | 13 | 2019 |
| 185 | Lund University | Sweden | 13 | 2015 |
| 186 | Kyushu University | Japan | 13 | 2017 |
| 187 | Johannes Gutenberg-University Mainz | Germany | 13 | 2013 |
| 188 | Huazhong Agricultural University | China | 13 | 2019 |
| 189 | China Pharmaceutical University | China | 13 | 2019 |
| 190 | Charite - University Medicine Berlin | Germany | 13 | 2011 |
| 191 | Beijing University of Chinese Medicine | China | 13 | 2015 |
| 192 | University of Virginia | USA | 12 | 2020 |
| 193 | University of Montreal | Canada | 12 | 2019 |
| 194 | University of Illinois | USA | 12 | 2019 |
| 195 | South China University of Technology | China | 12 | 2021 |
| 196 | Shaanxi University of Science and Technology | China | 12 | 2021 |
| 197 | Oregon State University | USA | 12 | 2016 |
| 198 | Nanchang University | China | 12 | 2019 |
| 199 | Jiangnan University | China | 12 | 2019 |
| 200 | Guangdong Medical University | China | 12 | 2017 |
| 201 | Ghent University | Belgium | 12 | 2020 |
| 202 | Baylor College of Medicine | USA | 12 | 2007 |
| 203 | Albert Einstein College of Medicine | USA | 12 | 2009 |
| 204 | Yokohama City University | Japan | 11 | 2018 |
| 205 | University of Paris | France | 11 | 2020 |
| 206 | University of Munster | Germany | 11 | 2010 |
| 207 | University of Massachusetts | USA | 11 | 2016 |
| 208 | University of London | England | 11 | 2016 |
| 209 | University of Alberta | Canada | 11 | 2011 |
| 210 | Umea University | Sweden | 11 | 2002 |
| 211 | Regensburg University Hospital | Germany | 11 | 2016 |
| 212 | Louisiana State University | USA | 11 | 2003 |
| 213 | Lincoln Memorial University | USA | 11 | 2015 |
| 214 | Jilin University | China | 11 | 2020 |
| 215 | Hebrew University of Jerusalmen | Israel | 11 | 2011 |
| 216 | Cleveland Clinic | USA | 11 | 2010 |
| 217 | Case Western Reserve University | USA | 11 | 2009 |
| 218 | Babraham Institute | England | 11 | 2009 |
| 219 | Autonomous University of Barcelona | Spain | 11 | 2017 |
| 220 | Zhejiang Chinese Medical University | China | 10 | 2020 |
| 221 | Xiamen University | China | 10 | 2021 |
| 222 | Wake Forest School of Medicine | USA | 10 | 2020 |
| 223 | University of Utah | USA | 10 | 2020 |
| 224 | University of Oslo | Norway | 10 | 2016 |
| 225 | University of Missouri | USA | 10 | 2007 |
| 226 | University of Liege | Belgium | 10 | 2019 |
| 227 | University of Graz | Austria | 10 | 2005 |
| 228 | Tianjin University | China | 10 | 2009 |
| 229 | SCIEX Ltd | USA | 10 | 2018 |
| 230 | Oregon Health and Science University | USA | 10 | 2017 |
| 231 | National Institute of Standards and Technology | USA | 10 | 2017 |
| 232 | Laval University | Canada | 10 | 2013 |
| 233 | Emory University | USA | 10 | 2019 |
| 234 | Chinese Academy of Medical Sciences and Peking Union Medical College | China | 10 | 2021 |

**Table S3**. The 425 keywords with more than 10 frequencies

| Rank | Keywords | Counts | **Time First** | Rank | Keywords | Counts | **Time First** |
| --- | --- | --- | --- | --- | --- | --- | --- |
| 1 | Mass Spectrometry | 1188 | 2005 | 214 | Degradation | 34 | 2017 |
| 2 | Metabolism | 962 | 2005 | 215 | Fatty Acid Metabolism | 33 | 2017 |
| 3 | Fatty Acid | 838 | 2002 | 216 | Diabetes Mellitus | 33 | 2014 |
| 4 | Identification | 757 | 2003 | 217 | Acyltransferase | 33 | 2011 |
| 5 | Expression | 565 | 2002 | 218 | Spectrometry | 33 | 2008 |
| 6 | Phospholipid | 461 | 2003 | 219 | Microalgae | 33 | 2014 |
| 7 | Shotgun Lipidomics | 449 | 2006 | 220 | Rat Brain | 33 | 2008 |
| 8 | Lipid | 449 | 2006 | 221 | Quality | 33 | 2018 |
| 9 | Lipidomics | 433 | 2006 | 222 | Biology | 33 | 2011 |
| 10 | Disease | 429 | 2004 | 223 | Insulin | 33 | 2011 |
| 11 | Protein | 399 | 2005 | 224 | Exosm | 33 | 2019 |
| 12 | Cell | 388 | 2004 | 225 | Diacylglycerol | 32 | 2004 |
| 13 | Insulin Resistance | 386 | 2007 | 226 | Pregnancy | 32 | 2019 |
| 14 | Oxidative Stress | 382 | 2007 | 227 | Steatosis | 32 | 2017 |
| 15 | Inflammation | 378 | 2002 | 228 | Er Stress | 32 | 2013 |
| 16 | Acid | 362 | 2003 | 229 | Fibrosis | 32 | 2016 |
| 17 | Tandem Mass Spectrometry | 347 | 2005 | 230 | Alpha | 32 | 2007 |
| 18 | Plasma | 341 | 2005 | 231 | Role | 32 | 2005 |
| 19 | Cholesterol | 317 | 2004 | 232 | Tolerance | 31 | 2014 |
| 20 | Electrospray Ionization | 308 | 2003 | 233 | Time | 31 | 2016 |
| 21 | Activation | 300 | 2003 | 234 | Oxidative Lipidomics | 30 | 2008 |
| 22 | Mechanism | 293 | 2007 | 235 | Involvement | 30 | 2018 |
| 23 | Risk | 292 | 2011 | 236 | Fatty Liver | 30 | 2010 |
| 24 | Gene Expression | 291 | 2007 | 237 | Population | 30 | 2018 |
| 25 | Metabolomics | 289 | 2007 | 238 | Modulation | 30 | 2017 |
| 26 | Biosynthesis | 281 | 2003 | 239 | Exercise | 30 | 2010 |
| 27 | Biomarker | 277 | 2009 | 240 | Product | 30 | 2015 |
| 28 | Lipid Metabolism | 271 | 2008 | 241 | Target | 30 | 2015 |
| 29 | Ceramide | 267 | 2005 | 242 | Sample | 30 | 2015 |
| 30 | Polyunsaturated Fatty Acid | 263 | 2005 | 243 | Heart | 30 | 2013 |
| 31 | Phosphatidylcholine | 263 | 2005 | 244 | Multidimensional Mass Spectrometry | 29 | 2010 |
| 32 | Liquid Chromatography | 250 | 2007 | 245 | Mild Cognitive Impairment | 29 | 2017 |
| 33 | Obesity | 240 | 2007 | 246 | Lipid Extraction | 29 | 2011 |
| 34 | Fatty Acid Composition | 234 | 2007 | 247 | Liver Disease | 29 | 2013 |
| 35 | Brain | 229 | 2003 | 248 | Proliferation | 29 | 2018 |
| 36 | Arachidonic Acid | 221 | 2005 | 249 | Platform | 29 | 2019 |
| 37 | Quantification | 215 | 2008 | 250 | Mass Spectrometric Analysis | 28 | 2008 |
| 38 | Docosahexaenoic Acid | 211 | 2002 | 251 | Chronic Kidney Disease | 28 | 2016 |
| 39 | Quantitative Analysis | 207 | 2004 | 252 | Lipid Profile | 28 | 2014 |
| 40 | Membrane | 203 | 2005 | 253 | Optimization | 28 | 2016 |
| 41 | Alzheimers Disease | 200 | 2003 | 254 | Cytochrome C | 28 | 2007 |
| 42 | Gene | 199 | 2007 | 255 | Synthase | 28 | 2010 |
| 43 | Growth | 197 | 2007 | 256 | Lipid Profiling | 27 | 2008 |
| 44 | Mice | 192 | 2009 | 257 | Generation | 27 | 2009 |
| 45 | Accumulation | 190 | 2006 | 258 | Disorder | 27 | 2014 |
| 46 | Cardiovascular Disease | 186 | 2004 | 259 | Domain | 27 | 2003 |
| 47 | Adipose Tissue | 181 | 2009 | 260 | Women | 27 | 2020 |
| 48 | Extraction | 180 | 2008 | 261 | Untargeted Lipidomics | 26 | 2018 |
| 49 | Biological Sample | 178 | 2003 | 262 | Energy Metabolism | 26 | 2014 |
| 50 | Inhibition | 174 | 2005 | 263 | Phospholipase D | 26 | 2007 |
| 51 | Cancer | 169 | 2006 | 264 | Heart Failure | 26 | 2017 |
| 52 | Receptor | 168 | 2006 | 265 | Omega-3 Fatty Acid | 25 | 2015 |
| 53 | Apoptosis | 166 | 2003 | 266 | Cholesteryl Ester | 25 | 2009 |
| 54 | Association | 165 | 2004 | 267 | Cellular Lipidm | 25 | 2005 |
| 55 | Pathway | 165 | 2007 | 268 | Validation | 25 | 2013 |
| 56 | Model | 161 | 2008 | 269 | Adaptation | 25 | 2014 |
| 57 | Serum | 153 | 2010 | 270 | Death | 25 | 2015 |
| 58 | Sphingolipid | 145 | 2008 | 271 | Lc M | 25 | 2017 |
| 59 | Stress | 145 | 2011 | 272 | Oxidized Phospholipid | 24 | 2007 |
| 60 | Membrane Lipid | 142 | 2005 | 273 | Targeted Lipidomics | 24 | 2005 |
| 61 | Tissue | 139 | 2009 | 274 | Sample Preparation | 24 | 2008 |
| 62 | Liver | 138 | 2008 | 275 | Type 2 Diabete | 24 | 2019 |
| 63 | Metabolic Syndrome | 132 | 2006 | 276 | Consumption | 24 | 2013 |
| 64 | Metabolite | 131 | 2006 | 277 | Eicosanoid | 24 | 2002 |
| 65 | Reveal | 130 | 2011 | 278 | Life Span | 24 | 2013 |
| 66 | Separation | 129 | 2006 | 279 | Induction | 24 | 2013 |
| 67 | Profile | 128 | 2007 | 280 | Lc Ms/M | 24 | 2017 |
| 68 | High Throughput | 119 | 2010 | 281 | Insight | 24 | 2017 |
| 69 | Plasma Membrane | 118 | 2005 | 282 | Unsaturated Fatty Acid | 23 | 2018 |
| 70 | In Vivo | 117 | 2007 | 283 | De Novo Lipogenesis | 23 | 2018 |
| 71 | System | 115 | 2010 | 284 | Apolipoprotein E | 23 | 2007 |
| 72 | Lipidomic Analysis | 114 | 2002 | 285 | Chlamydomonas Reinhardtii | 22 | 2015 |
| 73 | Performance Liquid Chromatography | 111 | 2005 | 286 | Chain Fatty Acid | 22 | 2012 |
| 74 | Resistance | 111 | 2011 | 287 | Systems Biology | 22 | 2008 |
| 75 | Phosphatidylethanolamine | 110 | 2011 | 288 | Human Brain | 22 | 2015 |
| 76 | Skeletal Muscle | 110 | 2010 | 289 | Cancer Cell | 22 | 2014 |
| 77 | Atherosclerosis | 104 | 2009 | 290 | Dementia | 22 | 2013 |
| 78 | Saccharomyces Cerevisiae | 102 | 2009 | 291 | Induced Insulin Resistance | 21 | 2012 |
| 79 | Human Plasma | 101 | 2012 | 292 | Alpha Linolenic Acid | 21 | 2014 |
| 80 | Risk Factor | 100 | 2011 | 293 | Barth Syndrome | 21 | 2008 |
| 81 | Resolution | 100 | 2009 | 294 | Prostaglandin | 21 | 2011 |
| 82 | Breast Cancer | 98 | 2010 | 295 | Polar Lipid | 21 | 2012 |
| 83 | Sphingomyelin | 98 | 2005 | 296 | Double Bond | 21 | 2019 |
| 84 | Quantitation | 98 | 2006 | 297 | Weight Lo | 21 | 2018 |
| 85 | Triacylglycerol | 95 | 2011 | 298 | Ester | 21 | 2013 |
| 86 | Lysophosphatidylcholine | 92 | 2008 | 299 | Non-Alcoholic Fatty Liver Disease | 20 | 2019 |
| 87 | Mouse Model | 91 | 2008 | 300 | Assisted Laser Desorption | 20 | 2007 |
| 88 | Structural Characterization | 90 | 2007 | 301 | Transcription Factor | 20 | 2015 |
| 89 | Hepatic Steatosis | 90 | 2007 | 302 | Blood Pressure | 20 | 2015 |
| 90 | Cardiolipin | 88 | 2007 | 303 | Trafficking | 20 | 2011 |
| 91 | Glucose | 88 | 2009 | 304 | Mellitus | 20 | 2016 |
| 92 | Chromatography | 87 | 2006 | 305 | Bacteria | 20 | 2015 |
| 93 | Health | 85 | 2007 | 306 | Choline | 20 | 2004 |
| 94 | Differentiation | 83 | 2012 | 307 | Muscle | 20 | 2014 |
| 95 | Rat | 82 | 2005 | 308 | Food | 20 | 2019 |
| 96 | Lipid Mediator | 81 | 2009 | 309 | Liquid Chromatography-Mass Spectrometry | 19 | 2015 |
| 97 | In Vitro | 81 | 2011 | 310 | Glycosphingolipid | 19 | 2008 |
| 98 | Ionization Mass Spectrometry | 80 | 2005 | 311 | Messenger Rna | 19 | 2015 |
| 99 | Low Density Lipoprotein | 80 | 2009 | 312 | Ganglioside | 19 | 2010 |
| 100 | Fatty Liver Disease | 79 | 2009 | 313 | Biogenesis | 19 | 2004 |
| 101 | Progression | 79 | 2015 | 314 | Adipocyte | 19 | 2011 |
| 102 | Endoplasmic Reticulum | 78 | 2003 | 315 | Lipolysis | 19 | 2019 |
| 103 | Arabidopsis | 77 | 2008 | 316 | Isomer | 19 | 2017 |
| 104 | Oxidation | 77 | 2011 | 317 | Fatty Acid Oxidation | 18 | 2017 |
| 105 | Insulin Sensitivity | 74 | 2010 | 318 | Signal Transduction | 18 | 2005 |
| 106 | Deficiency | 72 | 2007 | 319 | Metabolic Profiling | 18 | 2007 |
| 107 | Injury | 72 | 2011 | 320 | Machine Learning | 18 | 2020 |
| 108 | Lipid Peroxidation | 71 | 2005 | 321 | Susceptibility | 18 | 2019 |
| 109 | Phosphatidic Acid | 71 | 2007 | 322 | Lung Cancer | 18 | 2019 |
| 110 | Exposure | 71 | 2016 | 323 | Prevention | 18 | 2018 |
| 111 | Coronary Artery Disease | 70 | 2010 | 324 | Carcinoma | 18 | 2016 |
| 112 | Dysfunction | 70 | 2016 | 325 | Inhibitor | 18 | 2016 |
| 113 | Discovery | 69 | 2013 | 326 | Lipidome | 18 | 2020 |
| 114 | Lipid Composition | 68 | 2002 | 327 | Nonalcoholic Fatty Liver | 17 | 2018 |
| 115 | Crude Extract | 68 | 2005 | 328 | Beta Oxidation | 17 | 2012 |
| 116 | Endoplasmic Reticulum Stress | 67 | 2014 | 329 | Epidemiology | 17 | 2019 |
| 117 | Lipoprotein | 66 | 2008 | 330 | Elucidation | 17 | 2012 |
| 118 | Homeostasis | 64 | 2016 | 331 | Strain | 17 | 2015 |
| 119 | Fish Oil | 64 | 2010 | 332 | Kinase | 17 | 2018 |
| 120 | Arabidopsis Thaliana | 63 | 2006 | 333 | High-Resolution Mass Spectrometry | 16 | 2017 |
| 121 | Diagnosis | 63 | 2011 | 334 | Overexpression | 16 | 2019 |
| 122 | Escherichia Coli | 62 | 2007 | 335 | Ppar Alpha | 16 | 2014 |
| 123 | Temperature | 62 | 2014 | 336 | Increase | 16 | 2020 |
| 124 | Toxicity | 62 | 2016 | 337 | Ms/M | 16 | 2017 |
| 125 | Amino Acid | 61 | 2013 | 338 | Fish | 16 | 2002 |
| 126 | Response | 61 | 2014 | 339 | Nonalcoholic Fatty Liver Disease | 15 | 2016 |
| 127 | Plant | 61 | 2013 | 340 | Unfolded Protein Response | 15 | 2015 |
| 128 | Extracellular Vesicle | 60 | 2016 | 341 | Computational Lipidomics | 15 | 2020 |
| 129 | Mitochondria | 60 | 2011 | 342 | Collision Cross Section | 15 | 2015 |
| 130 | Cell Death | 60 | 2005 | 343 | Caenorhabditis Elegan | 15 | 2020 |
| 131 | Extract | 60 | 2006 | 344 | Biomarker Discovery | 15 | 2015 |
| 132 | Phospholipase A(2) | 59 | 2005 | 345 | Lipoprotein Lipase | 15 | 2015 |
| 133 | Omega 3 Fatty Acid | 59 | 2009 | 346 | Proteomic Analysis | 15 | 2009 |
| 134 | Blood | 59 | 2013 | 347 | Crystal Structure | 15 | 2003 |
| 135 | Fragmentation | 58 | 2011 | 348 | Cold Acclimation | 15 | 2015 |
| 136 | Pathogenesis | 58 | 2011 | 349 | Phosphorylation | 15 | 2019 |
| 137 | Lipid Droplet | 57 | 2014 | 350 | Organization | 15 | 2018 |
| 138 | Glycerophospholipid | 56 | 2011 | 351 | Force Field | 15 | 2019 |
| 139 | Ionization | 56 | 2006 | 352 | Antioxidant | 15 | 2015 |
| 140 | Database | 56 | 2008 | 353 | Dna Damage | 15 | 2015 |
| 141 | Eicosapentaenoic Acid | 55 | 2009 | 354 | Phenotype | 15 | 2013 |
| 142 | Peroxidation | 55 | 2012 | 355 | Droplet | 15 | 2019 |
| 143 | Mouse | 55 | 2010 | 356 | Storage | 15 | 2019 |
| 144 | Sphingosine 1 Phosphate | 54 | 2005 | 357 | Damage | 15 | 2013 |
| 145 | Sphingolipid Metabolism | 54 | 2009 | 358 | Tumor | 15 | 2017 |
| 146 | Survival | 54 | 2016 | 359 | Thin Layer Chromatography | 14 | 2008 |
| 147 | Enzyme | 54 | 2006 | 360 | Targeted Metabolomics | 14 | 2017 |
| 148 | Coronary Heart Disease | 53 | 2009 | 361 | Phosphatidylglycerol | 14 | 2018 |
| 149 | Colorectal Cancer | 53 | 2014 | 362 | Saturated Fatty Acid | 14 | 2012 |
| 150 | Free Fatty Acid | 53 | 2010 | 363 | Apolipoprotein A I | 14 | 2010 |
| 151 | Purification | 53 | 2008 | 364 | Parkinsons Disease | 14 | 2015 |
| 152 | Phosphatidylserine | 52 | 2007 | 365 | Epithelial Cell | 14 | 2005 |
| 153 | Plasmalogen | 52 | 2007 | 366 | Anandamide | 14 | 2008 |
| 154 | Triglyceride | 51 | 2011 | 367 | Management | 14 | 2020 |
| 155 | Performance | 51 | 2011 | 368 | Nafld | 14 | 2020 |
| 156 | Mediator | 51 | 2007 | 369 | Platelet Activating Factor | 13 | 2005 |
| 157 | Sensitivity | 50 | 2010 | 370 | Phospholipid Composition | 13 | 2008 |
| 158 | Macrophage | 50 | 2013 | 371 | Acid Sphingomyelinase | 13 | 2005 |
| 159 | Mutation | 50 | 2016 | 372 | Fatty Acid Synthesis | 13 | 2010 |
| 160 | Strategy | 50 | 2015 | 373 | Membrane Fluidity | 13 | 2019 |
| 161 | Diet | 50 | 2013 | 374 | Plasma Ceramide | 13 | 2019 |
| 162 | Yeast | 49 | 2010 | 375 | Deficient Mice | 13 | 2010 |
| 163 | Nonalcoholic Steatohepatiti | 48 | 2013 | 376 | High-Fat Diet | 13 | 2018 |
| 164 | Gas Chromatography | 48 | 2007 | 377 | Amyloid Beta | 13 | 2012 |
| 165 | Marker | 48 | 2017 | 378 | Abnormality | 13 | 2009 |
| 166 | Tool | 48 | 2009 | 379 | Gas Phase | 13 | 2009 |
| 167 | Chromatography Mass Spectrometry | 47 | 2010 | 380 | Esi Ms/M | 13 | 2008 |
| 168 | High Density Lipoprotein | 47 | 2010 | 381 | Outcm | 13 | 2020 |
| 169 | Impact | 47 | 2019 | 382 | Dha | 13 | 2020 |
| 170 | Binding | 46 | 2003 | 383 | Membrane Phospholipid | 12 | 2008 |
| 171 | Mitochondrial Dysfunction | 45 | 2007 | 384 | Molecular Mechanism | 12 | 2019 |
| 172 | Lysophosphatidic Acid | 45 | 2005 | 385 | Cardiovascular Risk | 12 | 2016 |
| 173 | Cerebrospinal Fluid | 45 | 2007 | 386 | Fatty Acid Profile | 12 | 2020 |
| 174 | Protein Kinase C | 45 | 2011 | 387 | Mitochondrial | 12 | 2019 |
| 175 | Localization | 45 | 2009 | 388 | Air Pollution | 12 | 2020 |
| 176 | Proteomics | 45 | 2010 | 389 | Double Blind | 12 | 2009 |
| 177 | Double Bond Position | 44 | 2010 | 390 | Efficacy | 12 | 2019 |
| 178 | Supplementation | 44 | 2016 | 391 | Therapy | 12 | 2015 |
| 179 | Prediction | 44 | 2017 | 392 | Phase | 12 | 2017 |
| 180 | Prevalence | 44 | 2015 | 393 | Raft | 12 | 2013 |
| 181 | Infection | 44 | 2014 | 394 | Electrospray Ionization Mass Spectrometry | 11 | 2003 |
| 182 | Steatohepatiti | 43 | 2011 | 395 | Resolution Mass Spectrometry | 11 | 2017 |
| 183 | Gut Microbiota | 43 | 2017 | 396 | Ion Mobility Spectrometry | 11 | 2020 |
| 184 | Bile Acid | 43 | 2007 | 397 | Lipid Accumulation | 11 | 2017 |
| 185 | Diversity | 42 | 2009 | 398 | Freezing Tolerance | 11 | 2019 |
| 186 | Mortality | 42 | 2015 | 399 | Multiple Sclerosis | 11 | 2019 |
| 187 | Myocardial Infarction | 41 | 2009 | 400 | Induced Apoptosis | 11 | 2008 |
| 188 | Animal Model | 41 | 2015 | 401 | Immune Response | 11 | 2016 |
| 189 | Lipid Raft | 41 | 2005 | 402 | Alpha Synuclein | 11 | 2015 |
| 190 | Age | 41 | 2012 | 403 | Gut Microbiome | 11 | 2019 |
| 191 | Fat | 41 | 2009 | 404 | Heart Disease | 11 | 2014 |
| 192 | Secretion | 40 | 2015 | 405 | Metaanalysis | 11 | 2020 |
| 193 | Autophagy | 40 | 2018 | 406 | Stability | 11 | 2019 |
| 194 | Oil | 40 | 2015 | 407 | Component | 11 | 2010 |
| 195 | Imaging Mass Spectrometry | 38 | 2011 | 408 | Longevity | 11 | 2014 |
| 196 | Ozone Induced Dissociation | 37 | 2017 | 409 | Complex | 11 | 2015 |
| 197 | Mass Spectrometry Imaging | 37 | 2014 | 410 | Matrix | 11 | 2011 |
| 198 | Hepatocellular Carcinoma | 37 | 2015 | 411 | Lipidomic Profile | 10 | 2019 |
| 199 | Prostate Cancer | 37 | 2011 | 412 | Body Mass Index | 10 | 2019 |
| 200 | Ppar Gamma | 37 | 2016 | 413 | Cytochrome P450 | 10 | 2014 |
| 201 | Transport | 37 | 2016 | 414 | Infant Formula | 10 | 2020 |
| 202 | Release | 37 | 2007 | 415 | Human Meibum | 10 | 2014 |
| 203 | Ma | 37 | 2019 | 416 | Metabonomics | 10 | 2015 |
| 204 | High Throughput Quantification | 36 | 2009 | 417 | White Matter | 10 | 2017 |
| 205 | Linoleic Acid | 36 | 2013 | 418 | Bisphenol A | 10 | 2019 |
| 206 | Children | 36 | 2013 | 419 | Oleic Acid | 10 | 2015 |
| 207 | Fatty Acid Synthase | 35 | 2015 | 420 | Annotation | 10 | 2019 |
| 208 | Endothelial Cell | 35 | 2002 | 421 | Stem Cell | 10 | 2016 |
| 209 | Blood Plasma | 35 | 2012 | 422 | Vitamin E | 10 | 2016 |
| 210 | Dynamics | 35 | 2014 | 423 | Delivery | 10 | 2019 |
| 211 | Central Nervous System | 34 | 2007 | 424 | Mutant | 10 | 2007 |
| 212 | Multiple Precursor | 34 | 2007 | 425 | Drug | 10 | 2018 |
| 213 | Classification | 34 | 2015 |  |  |  |  |
